# Suitability of Google Trends™ for digital surveillance during ongoing COVID-19 epidemic: a case study from India

**DOI:** 10.1101/2020.08.24.20176321

**Authors:** Parmeshwar Satpathy, Sanjeev Kumar, Pankaj Prasad

**Affiliations:** Department of Community & Family Medicine, All India Institute of Medical Sciences (AIIMS), Bhopal, Madhya Pradesh, India

**Keywords:** disease surveillance, infodemiology, ICT in healthcare, Pandemic, time lag correlation

## Abstract

**Background:** India went into the largest population-level lockdown on 25^th^ March 2020 in response to the declaration of COVID-19 pandemic by World Health Organization (WHO). Digital surveillance has been shown to be useful to supplement the traditional surveillance. Google Trends™ (GT) is one such platform reported to be useful during pandemics of H1N1, Ebola and MERS.

**Objective:** We used GT to correlate the information seeking behaviour regarding COVID-19 of Indians with curiosity and apprehensiveness generated through media coverage as well as status of the epidemic both at national and state levels.

**Methods:** We retrieved GT data between 1^st^ January 2020 to 31^st^ May 2020 for India using a comprehensive search strategy. We obtained data on daily tests and cases from WHO, ECDC and covid19india.org websites. We explored the trends of COVID-19 in the form of relative search volume (RSV) from GT platform and correlated them with media reports. We used time-lag correlation analysis to assess the temporal relationships between Google search terms and daily new COVID-19 cases and daily tests for 14 days.

**Results:** Peaks in RSV correlated with media coverage or government declarations suggestive of curiosity and apprehensiveness both at national level and high-burden states. High time-lag correlation was observed between both the daily reported number of tests and cases and RSV for the terms “COVID 19”, “COVID”, “social distancing”, “soap” and “lockdown” at national level. Similar high time-lag correlation was observed for the terms “COVID 19”, “COVID”, “Corona”, “social distancing”, “soap”, “lockdown” in five high-burden states.

**Conclusion:** This study reveals the advantages of infodemiology using GT to monitor an emerging infectious disease like COVID-19 in India. Google searches in India during the ongoing COVID-19 pandemic reflects mostly curiosity and apprehension of citizens. GT can also complement traditional surveillance in India as well as high burden states.

## Introduction

With more than 6.6 million cases and more than 0.37 million deaths reported worldwide as on June 1^st^ 2020; the Covid-19 Pandemic is the biggest threat humanity has ever seen in over a century of the deadly Influenza Pandemic (Spanish Flu) of 1918. (1,2) According to WHO, COVID-19 is ten times as dangerous than Influenza.(3) Although presently healthcare facilities are being overwhelmed by COVID-19 in many countries, the situation was even worse in 1918, as hospitals were also dealing with mass casualties and injuries from the first world war.(4) Countries across the globe have been trying various pharmacological and non-pharmacological measures to combat this deadly pandemic.

India too did come up with various containment and mitigation measures. The country went into the largest population-level lockdown from 25^th^ March 2020 till 31^st^ May 2020 in response to the declaration of SARS-CoV-2 pandemic by World Health Organization (WHO) on 11^th^ March 2020. (5) The federal structure of the India leads to division of epidemic control responsibilities between union and state governments. Prior to the country-wide lockdown, the Union government first issued advisory to avoid travel to China followed by thermal screening of all international inbound passengers in early February 2020. Some of the states actively followed-up the international travelers, quarantined them or admitted them in hospitals, even before national level lockdown. Subsequently, the disease surveillance mechanism got activated, especially the Integrated Disease Surveillance Project (IDSP) network at state level through deployment of rapid response teams (RRTs).

Major uses of disease surveillance in the settings of epidemics are containment and control, risk communication and mitigation strategies for future similar events. In traditional disease surveillance, designated health staff or non-health trained personnel obtains information on any unusual disease activity. India’s traditional disease surveillance mechanism under IDSP relies on collection of data on diseases reported on weekly basis by health workers, doctors and laboratories in the country. (ref) Such weekly reporting of surveillance data can’t help in obtaining information for action related to containment and control during epidemics. There have been efforts to expedite the analysis of surveillance data through Integrated Health Information Platform (IHIP) of government of India which was initially launched to support IDSP in selected districts. Moreover, nearly 80% of ambulatory healthcare in India is provided by private health system and information in IDSP surveillance network is mostly of public system origin. Hence, the quality of traditional disease surveillance in India needs supplementation by other modes. There are various instances of use of Information, Communication & Technology (ICT) methods such as internet in disease surveillance. ‘Digital surveillance’ has been used during the previous H1N1, Ebola, Zika, Chikungunya outbreaks with varied results.(6) With over 560 million internet users, India ranks second in the world in terms of internet users; the first being China.(7) So a major portion of India’s population are internet users who in times of pandemic and the resulting lockdown will resort to the internet to find all possible information related to the COVID-19 pandemic. Risk communication to susceptible population is another requirement during epidemics to balance the knowledge and behaviour of the community to avoid both panic and indifference. Perception of hazard by community is one of the important factors in designing risk communication strategy. Based on high perceived susceptibility, people may resort to obtain information related to a particular disease. Surveillance of this information seeking behaviour can supplement disease surveillance. Researchers have analyzed the digital platforms like Google™ Search Engine, Twitter™, YouTube™ etc to understand the information seeking behaviour and predict its relationship with the disease status at population level. Google is most commonly used search Engine globally for information seeking including those for health conditions. One of the tool that allows users to analyze Internet search data is the freely accessible online portal known as Google Trends™ (GT). (8,9) Google Trends™ is an automated platform which provides analysis of Google search queries expressed in form of Relative Search Volume (RSV). It has been used to determine information seeking related to curiosity as well as prediction of cases during epidemics prone diseases such as H1N1 influenza, Ebola Virus Disease, Dengue Fever, Zika, MERS, conjunctivitis etc as well as for other diseases such as cancer, screening, Sinusitis, scarlet fever (10–23) A study from India reported time-lag correlation between GT and IDSP reporting by 1-3 weeks. Information seeking on SARS-CoV-2 can similarly be used for digital surveillance and inform policy making for containment and mitigation of COVID-19. During the current COVID-19 pandemic too, many studies have been published showing usefulness of GT.(24–33) Most of these studies have not used comprehensive search strategy and used few COVID-19 related terms.(29) We could identify only one study which included search trend data from India.(34) But this study reported search trend till initial 15 days of lockdown and used only one search term “COVID-19”. For supplementing infodemiological evidence, GT based data using comprehensive search strategy both at national and state level would be required.

Therefore, this study explored the potential use of Google Trends™ (GT) to monitor interest and apprehension of the population towards the COVID-19 pandemic in India. We also intended to explore potential of Google Trends™ as a tool in digital surveillance at level of country and states through determination of correlation between search behaviour and reported tests and cases. We wanted to explore whether such digital surveillance can be used to make future predictions during an outbreak or epidemic situation like current COVID-19 pandemic.

## Methods

### Data on COVID-19 cases and deaths

We accessed websites of the Ministry of Health and Family Welfare (MoHFW) of Government of India, the website covid19india.org, WHO COVID database and from European Centre for Disease Control (ECDC) to obtain data on COVID-19 tests, cases and deaths in India.(1,35–37) WHO COVID-19 database and ECDC had data on daily cases and deaths at national level. We could find data on tests, cases and deaths on a daily basis on the covid19india.org website both for country and state-levels. The data on WHO COVID-19 dashboard and ECDC were same for India. Data on covid19india.org was different from WHO COVID-19 dashboard. MoHFW had data only on the total number of cases and deaths on a particular day. It didn’t provide data of tests or cases or death from the previous days. We decided to use the data available on WHO COVID-19 dashboard for cases in India. Since data on tests and cases (state-level) were not available here, we decided to use covid19india.org for these data.

### Google Trends™ data on COVID-19 related search behaviour

We explored Google Trends™ between 3^rd^ and 4^th^ June 2020 to obtain search trends between 1^st^ January, 2020 (WHO got the information about possible novel Coronavirus outbreak on 31^st^ December) to May 31^st^, 2020 (Last day of lockdown in India).

We developed a list of 88 terms related to COVID- 19 both in English and Hindi languages (Table 1). These search terms represent the information search related for COVID-19 by people on google in India. We used both Roman and Unicode scripts for the Hindi terms. We then grouped these terms into five categories namely disease/ organism related terms; non-pharmacological interventions; symptoms; government instructions and all COVID related terms in Hindi language. We initially explored individual terms for trend during the study period. We couldn’t find any trend for 21 terms and excluded them. We further conducted Google Trend search of all the terms together (maximum five at a time which was permitted) from the corresponding group. In this stage, we had to remove another 49 terms which did not have trends at their group level. Finally, we had 14 terms in English and four terms in Hindi for our Google Trend search.

**Table 1:**
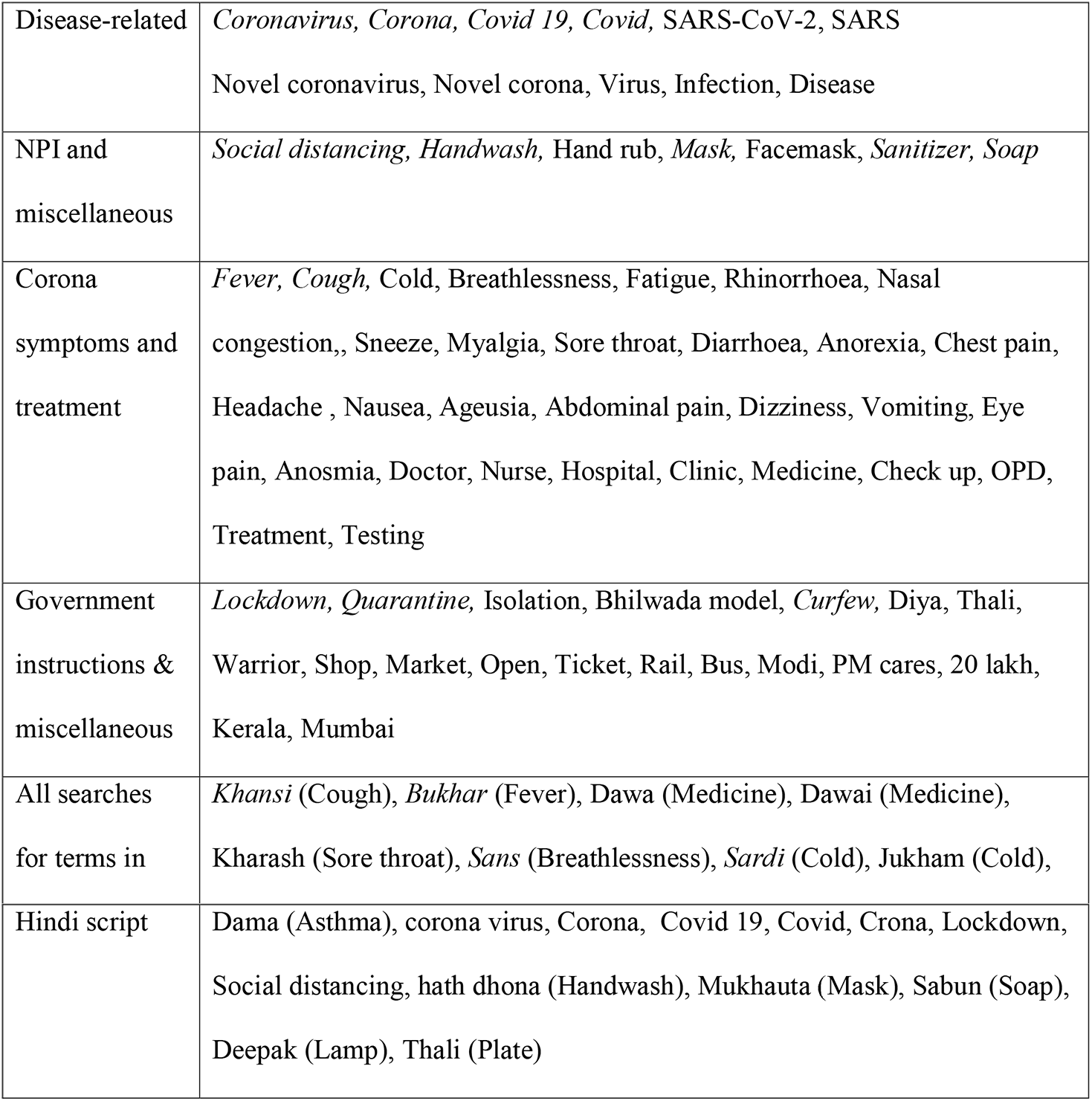
Search Strategy on Google Trends of Covid 19 in India

We filtered the Relative search volume (RSV) data both at national level as well as by geographic regions (states and Union Territories) of India. For the later, we included five states and union territories (UTs) each having high and low caseloads on 4^th^ June 2020. State/ UTs with high caseloads were Maharashtra, Rajasthan, Gujarat, Tamil Nadu and Delhi. States/UTs with low caseloads were Arunachal Pradesh, Daman Diu including Dadra Nagar Haveli, Mizoram, Sikkim, Meghalaya. Search data from the Google Trend website were exported in .csv format and later converted into .xlsx format for analysis. The results were obtained in form of graphs and tables. RSV of ‘100’ suggested maximum search interest while ‘0’ suggested no search interest for a particular term.

### Correlation between Google Trends™ data with daily tests and cases

We compared the Google Trends™ (GT) data with daily data on COVID-19 tests and cases both at national and the ten selected state levels. We used Pearson’s correlation analysis to examine the correlations of RSV data of Google search terms with daily tests conducted and daily new laboratory confirmed COVID-19 cases separately. We used the advanced data analysis tools available in Microsoft Excel 365 for this. We considered the correlation coefficient (r) of ≥ 0.5 to shows correlation between the search terms and daily count of tests or cases. We used time lag correlation analysis to assess the temporal relationships between these data for up to 14 days. The level of significance was set at 95%. We also conducted sensitivity analysis with data on cases and deaths between covid19india.org and ECDC data.

## Results

### Summary of testing and reported cases

Till 31^st^ May 2020, 6,028,326 cases and 368,944 deaths of Covid-19 were reported globally according to WHO COVID database and ECDC.(1,37) We could not find any record of daily and cumulative number of tests conducted at global level in any of the above databases. The corresponding numbers for India were 3,737,027 tests 182,143 cases and 5164 deaths according to Press Information Bureau of India (PIB) and ECDC database.(38,37) The daily tests peaked on 29^th^ May 2020 with 127,761 tests for India.(36) Testing capacity has consistently increased in India. The reported number of tests was 6000 on 13^th^ March and crossed to 100,000 on 18^th^ May 2020.(36) The first case from India was reported on 30^th^ January 2020 followed by one more case each on 2^nd^ and 3^rd^ February 2020, all from Kerala state. After a lull of nearly one month, two cases were reported on 2^nd^ March 2020, one each from Delhi Union Territory and Telangana state respectively. Daily new confirmed COVID-19 cases started to increase steadily ever since, with highest reported to be 8380 (WHO, ECDC and PIB) and 8789 on covid19india.org website on 31^st^ May 2020. First death from India was reported on 10^th^ March from Karnataka state.(39) There were 74 cases reported till this time. Similar to the consistent increase in cases, deaths due to COVID-19 has shown increasing trend, with 265 deaths reported on 30^th^ May 2020.

Till 31^st^ May 2020, highest number of cumulative cases were reported from the states/ UTs of Maharashtra, Tamil Nadu, Delhi, Gujarat and Rajasthan which was 67655, 22333, 19844, 16794 and 8831 respectively. Lowest cumulative cases were reported by states/UTs of Sikkim, Mizoram, Daman & Diu, Arunachal Pradesh and Meghalaya which stands at 1, 1, 2, 4 and 27 cases respectively. The number of cumulative tests for the five states/ UTs with highest caseloads were 463177, 491962, 212784, 211930 and 409777 respectively. The number of cumulative tests for the states/ UTs with the lowest caseloads were 2985, 777, 11477, 8283 and 7781 respectively on 31^st^ May 2020.

### Google Trend analysis for COVID- 19 related search behaviour

Figures 1 and 2 provide visual description of trends of the 18 terms from five groups on Google Trends™ at national level. Figures 3a to 7 depicts the similar trends for the 10 states included for analysis. In these figures, the left y-axis shows RSV values (0-100), while the right y-axis (secondary axis) shows either daily tests conducted (at national or state levels) or daily cases reported (at national or state levels). On the x-axis, the data interval is of one day. Figure 8 to 12 depicts time-lag correlation between the Google Trends ™ and daily tests or cases at the state level. There are two to five curves representing the trends in these figures. There are four tables (Tables 2-5) which are self-explanatory.

**Fig 1:**
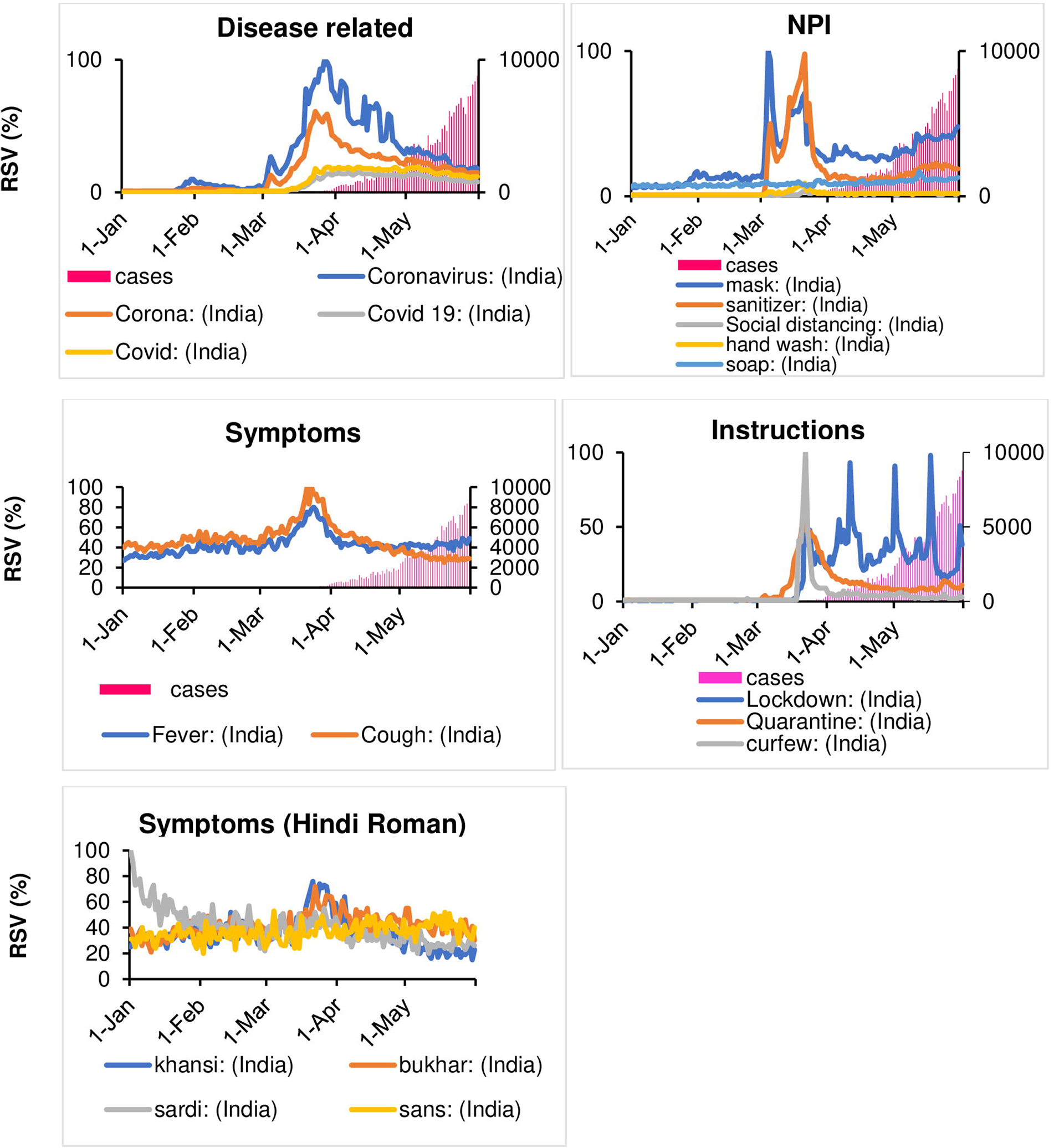
Time series of Google relative search volumes (RSVs) related to COVID-19 related search terms and daily COVID-19 cases in India

**Fig 2:**
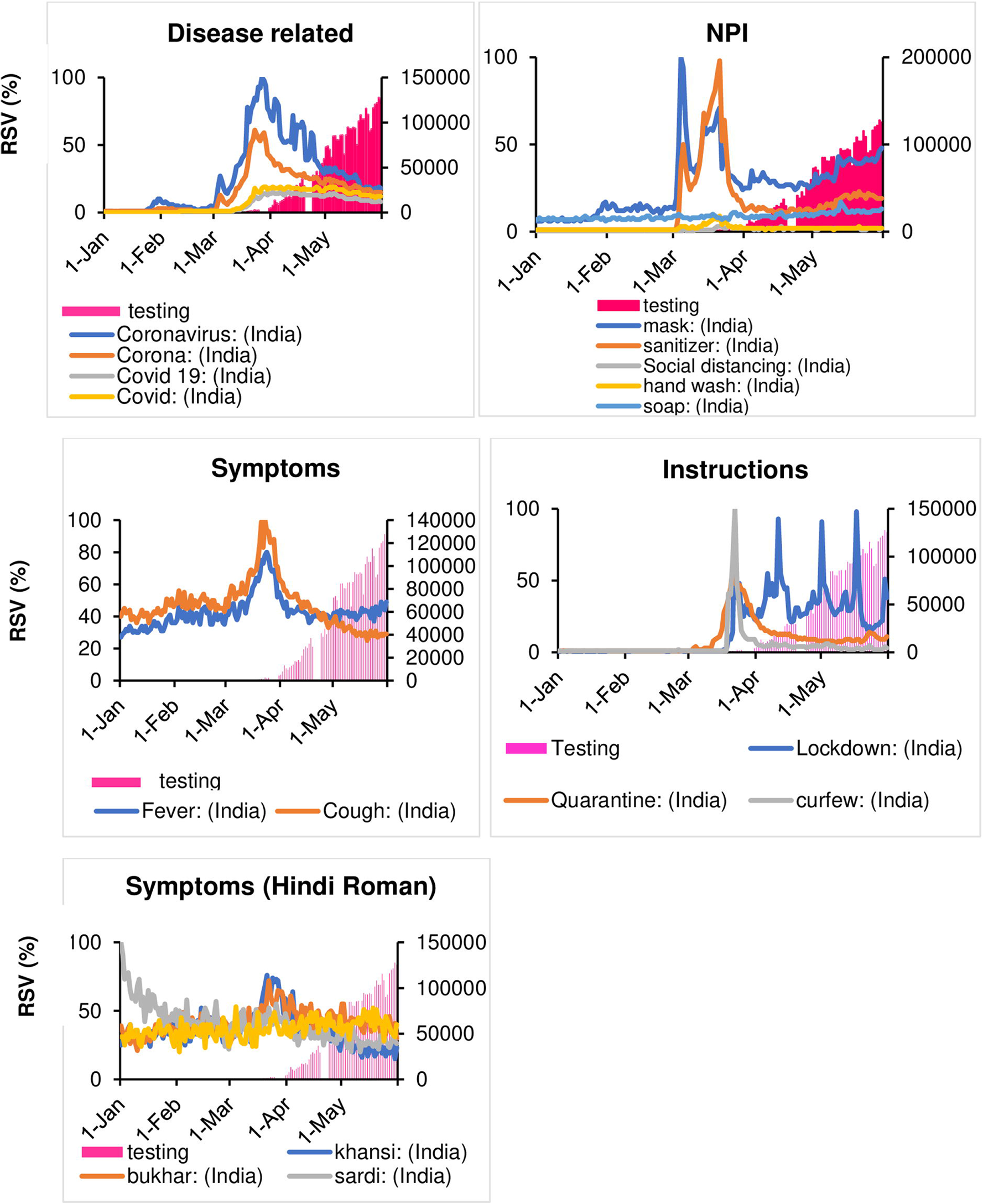
Time series of Google relative search volumes (RSVs) related to COVID-19 related search terms and daily COVID-19 testing in India

**Fig 3a:**
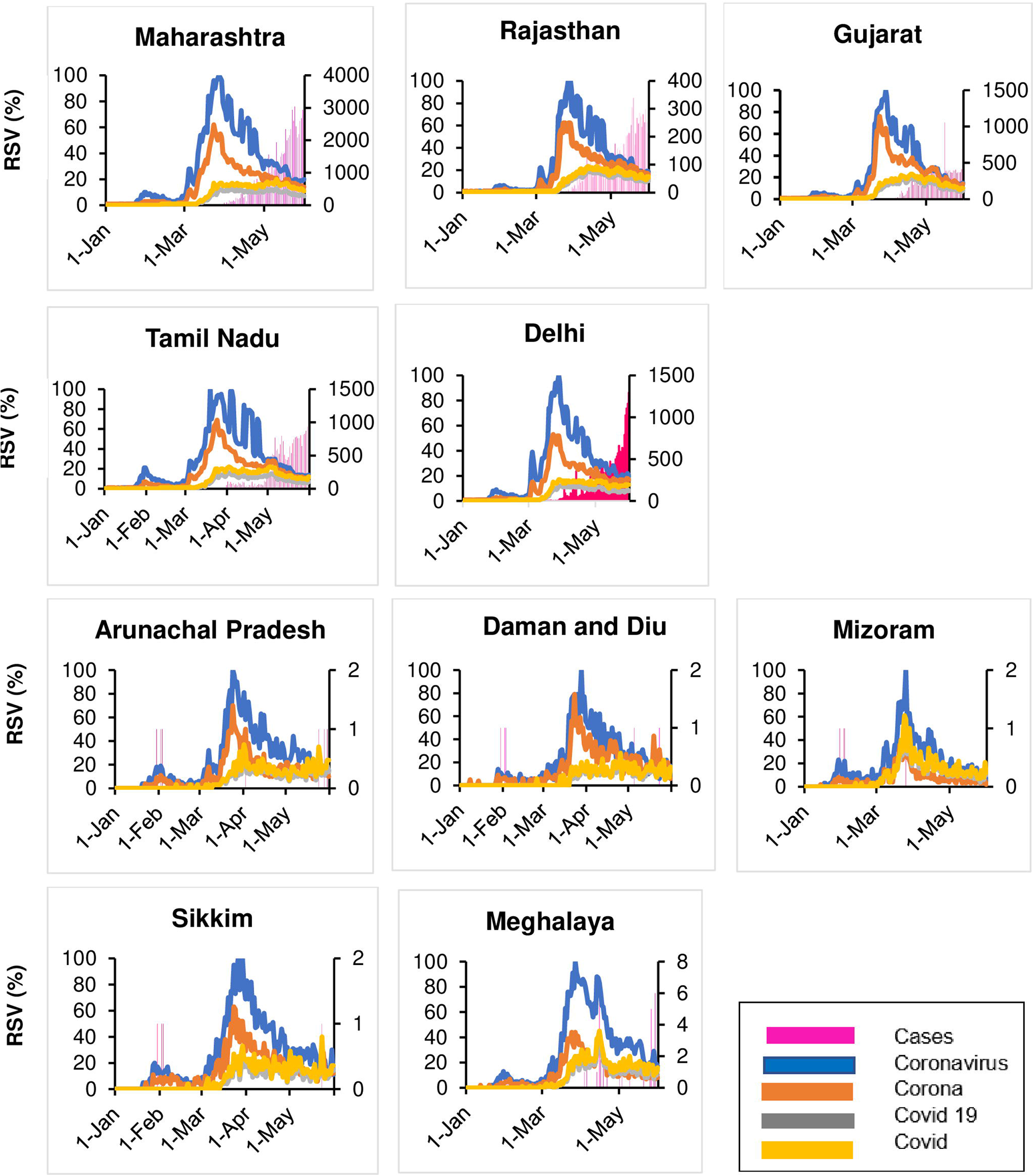
Time Series of Google Relative Search Volume (RSV) for COVID-19 disease related search terms and newly confirmed COVID-19 cases for selected states under study

**Fig 3b:**
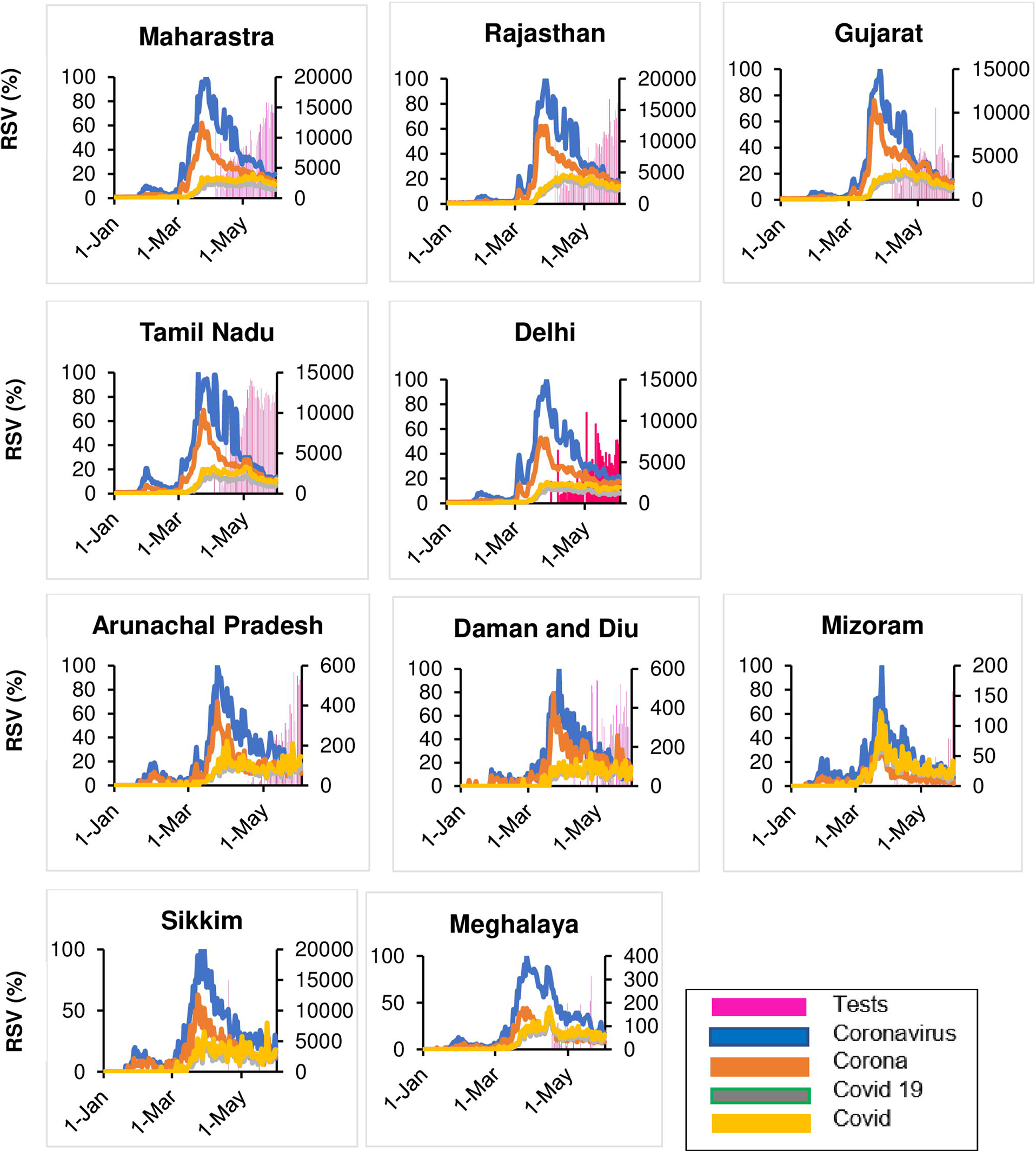
Time Series of Google Relative Search Volume (RSV) for COVID-19 disease related search terms and daily COVID-19 testing for selected states under study

**Fig 4a:**
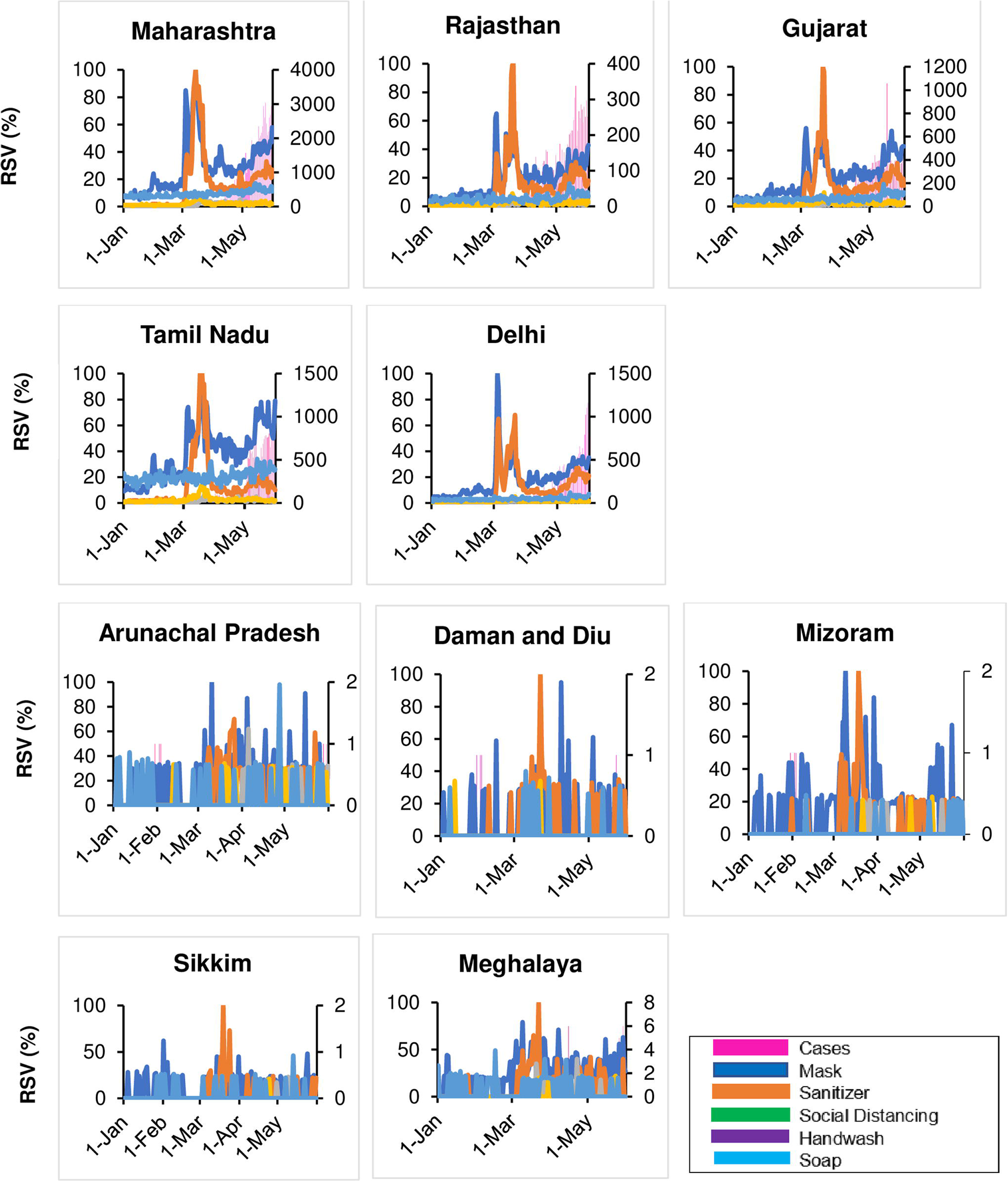
Time Series of Google Relative Search Volume (RSV) for NPI related search terms and newly confirmed COVID-19 cases for selected states under study

**Fig 4b:**
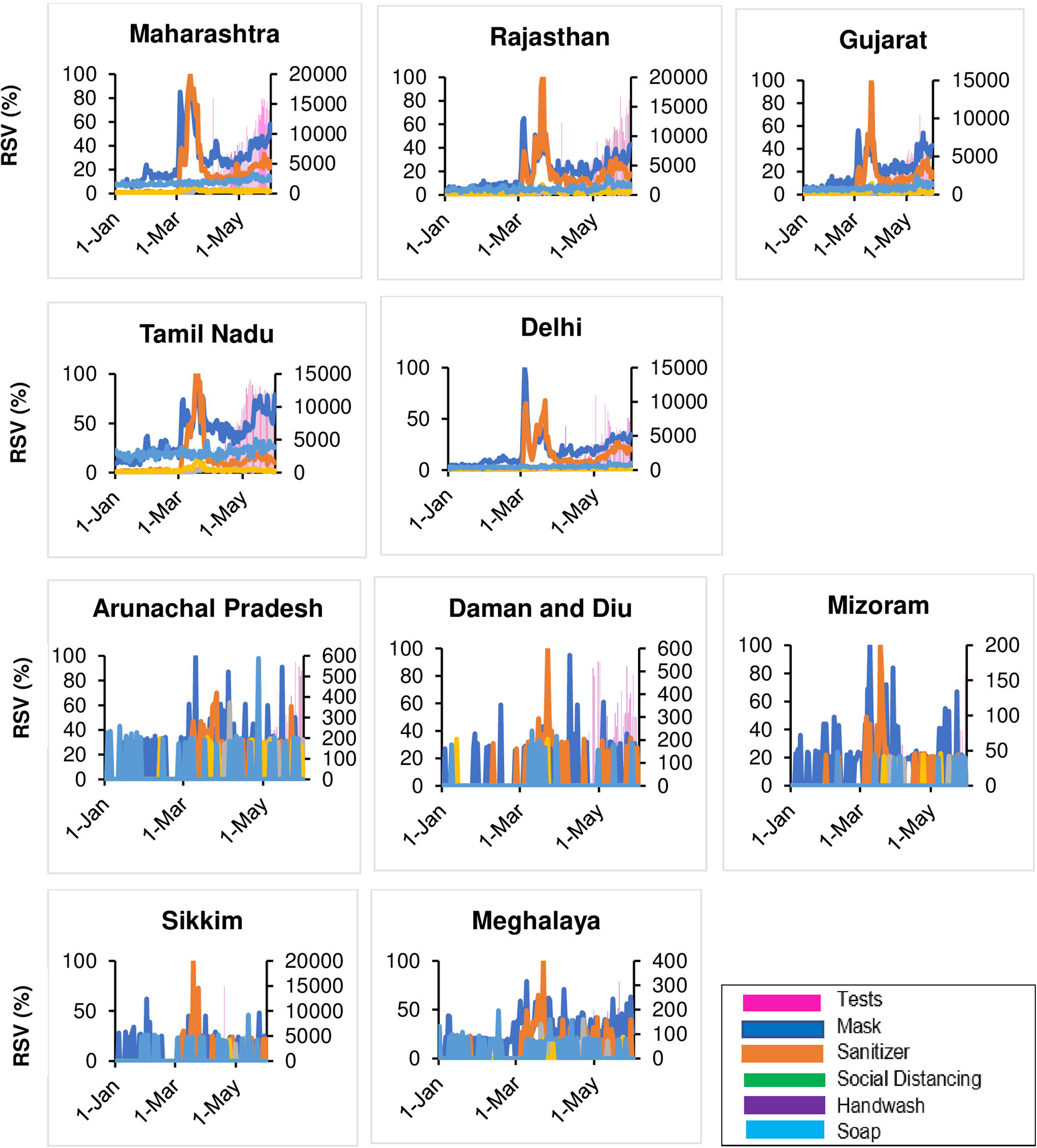
Time Series of Google Relative Search Volume (RSV) for NPI related search terms and daily COVID-19 tests for selected states under study

**Fig 5a:**
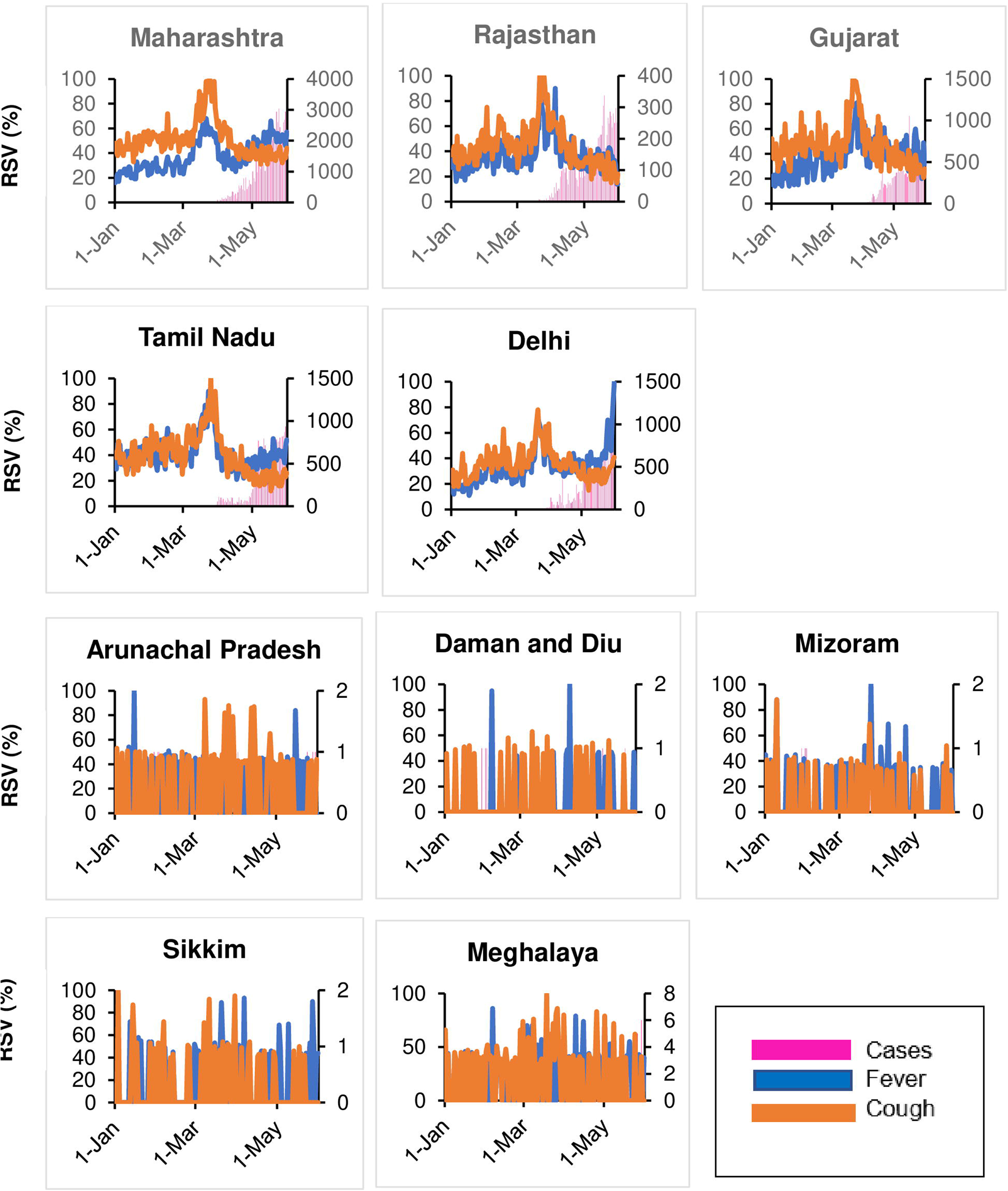
Time Series of Google Relative Search Volume (RSV) for COVID-19 symptoms and newly confirmed COVID-19 cases for selected states under study

**Fig 5b:**
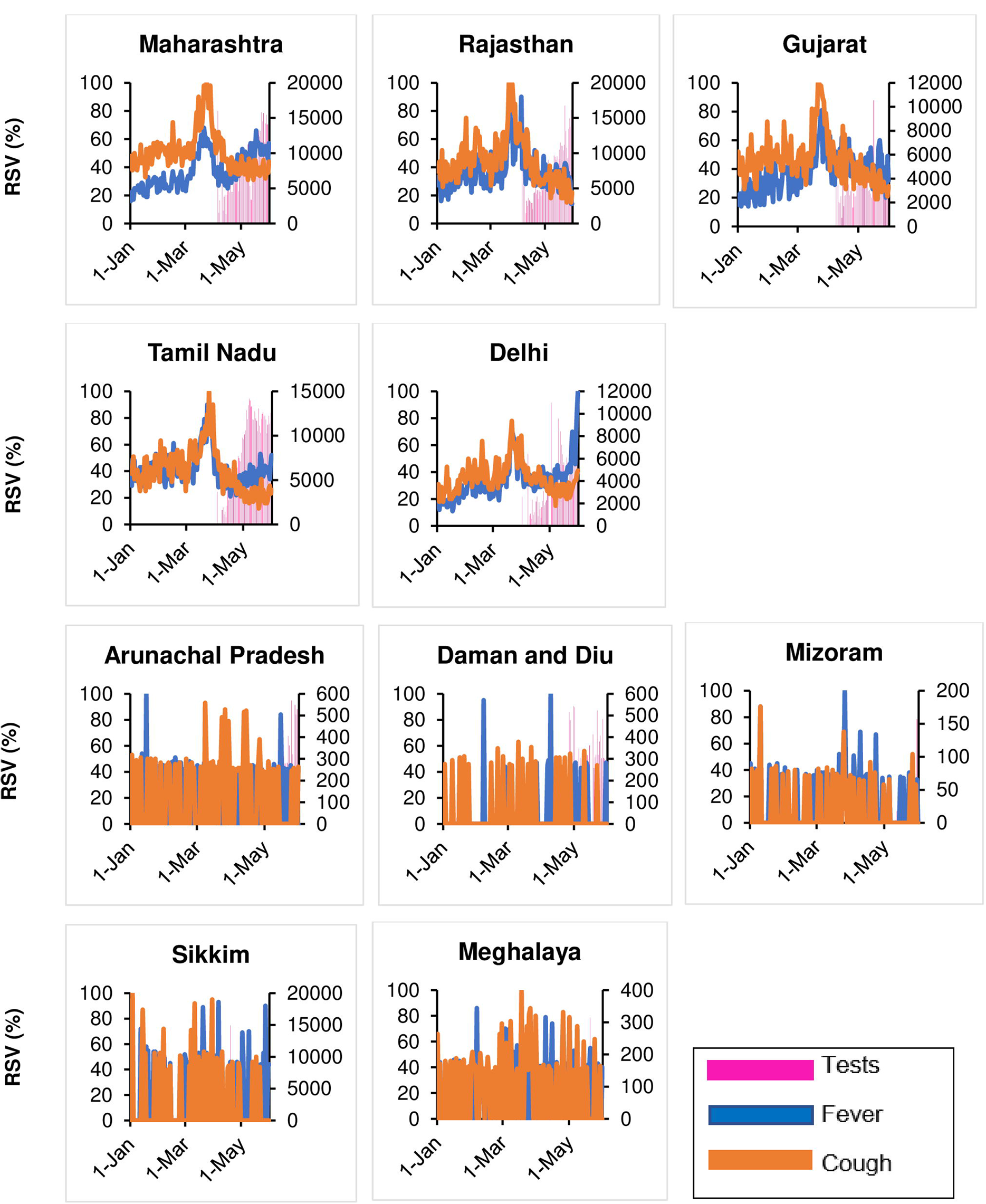
Time Series of Google Relative Search Volume (RSV) for COVID-19 symptoms and daily tests for selected states under study

**Fig 6a:**
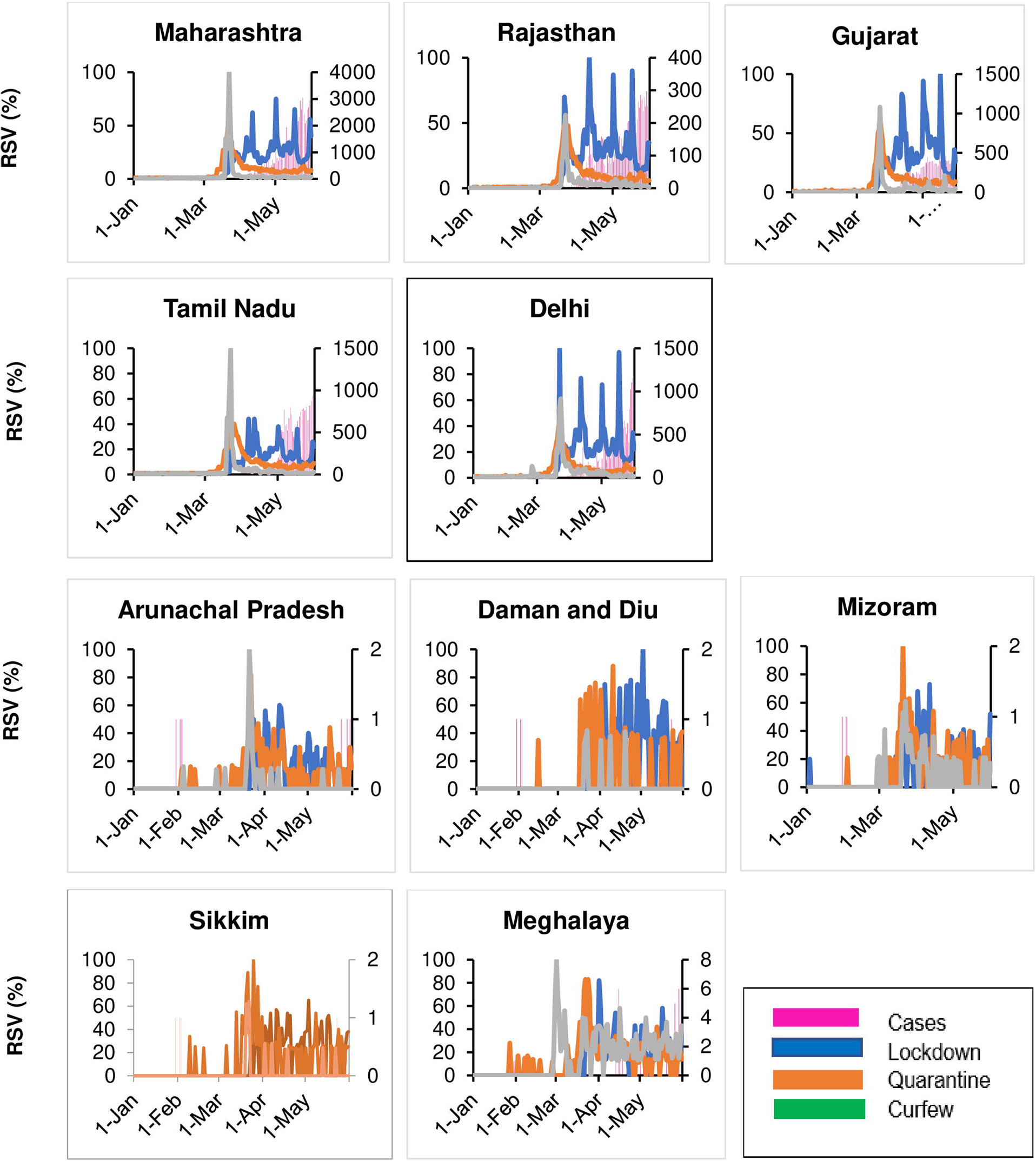
Time Series of Google Relative Search Volume (RSV) for Government instructions and newly confirmed COVID-19 cases for selected states under study

**Fig 6b:**
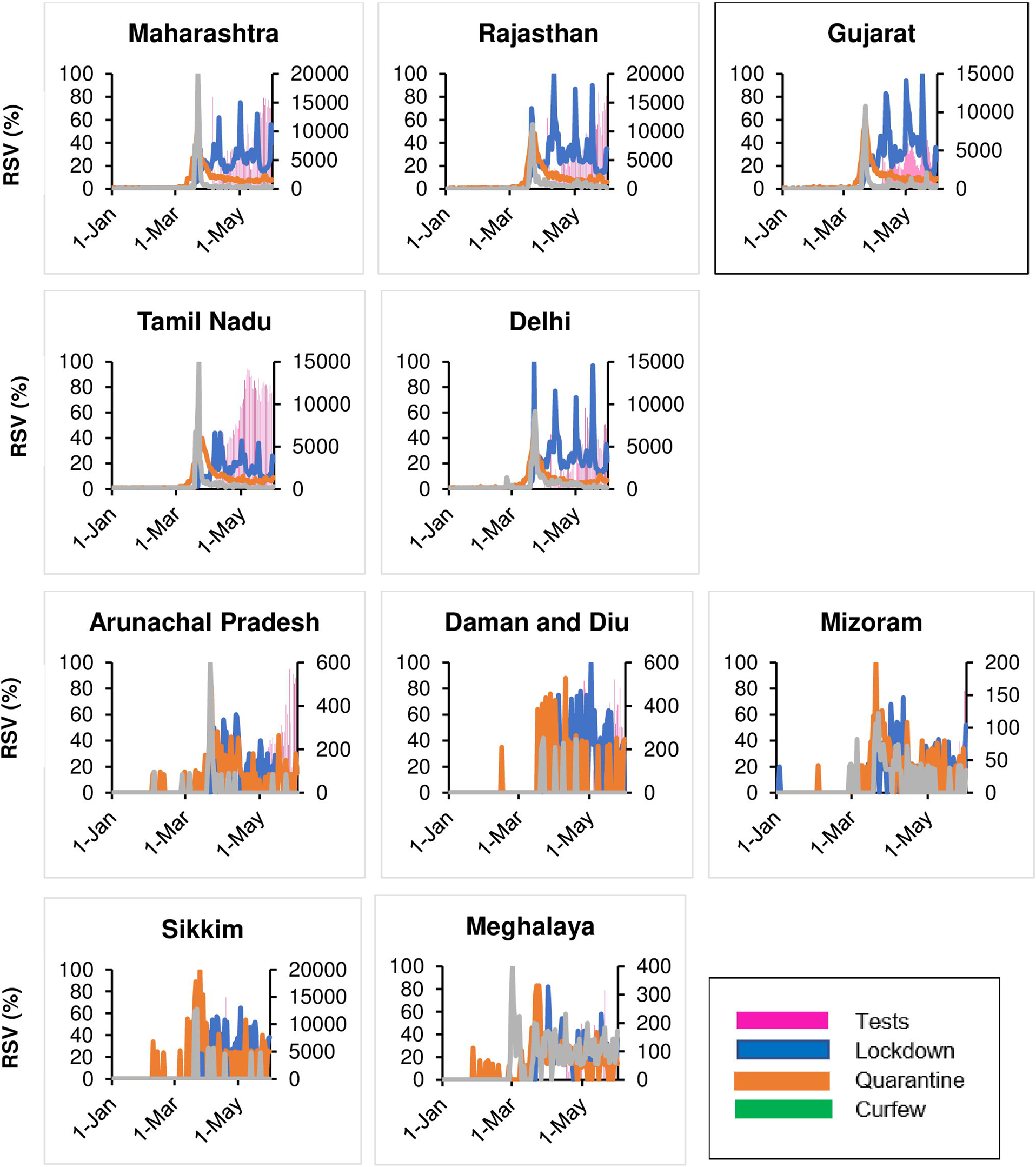
Time Series of Google Relative Search Volume (RSV) for Government instructions and daily new tests for selected states

**Fig 7:**
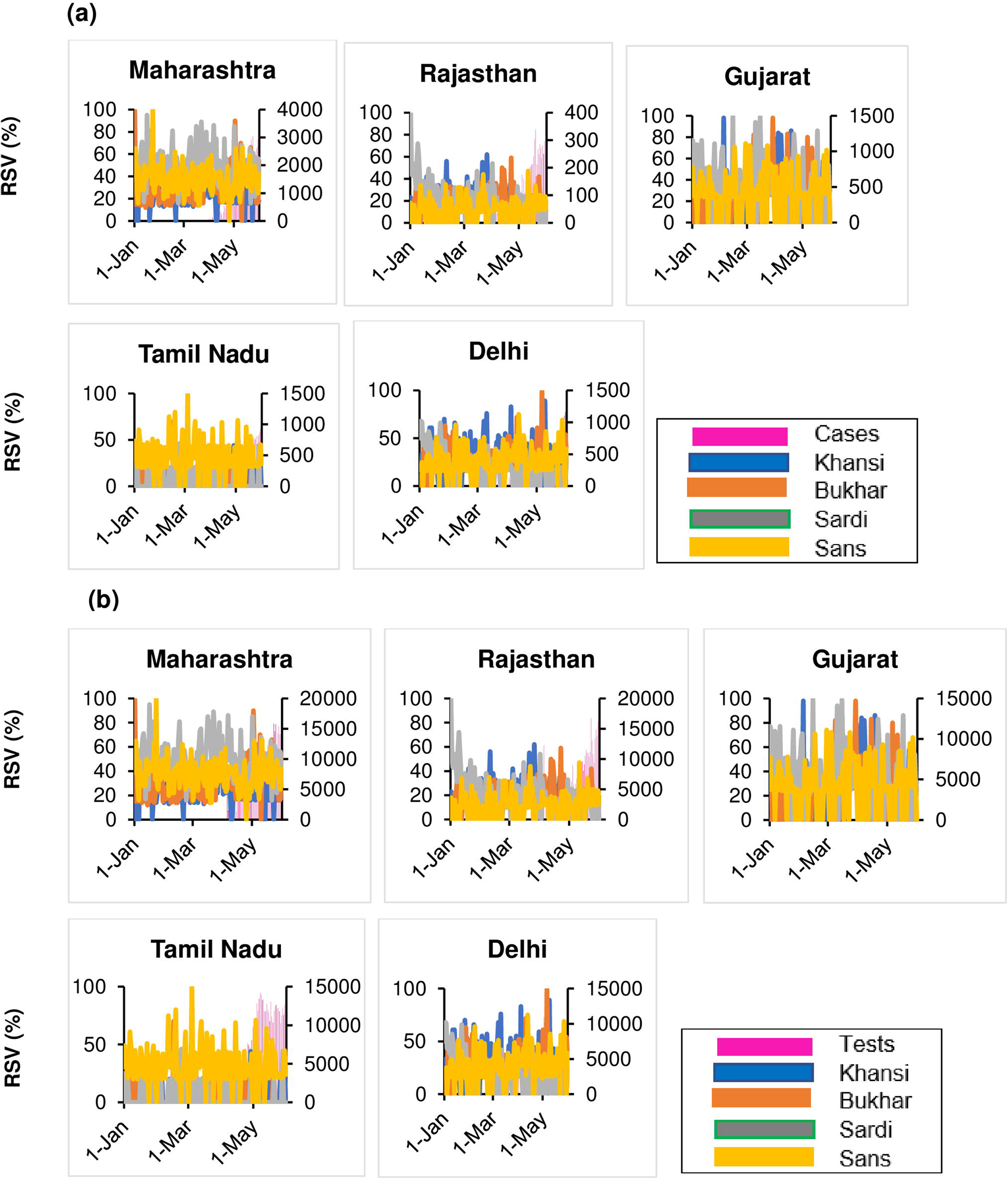
Time Series of Google Relative Search Volume (RSV) for COVID-19 symptoms in Hindi (Roman script) with (a) newly confirmed COVID-19 cases and (b) daily new tests for selected states under study

**Fig 8:**
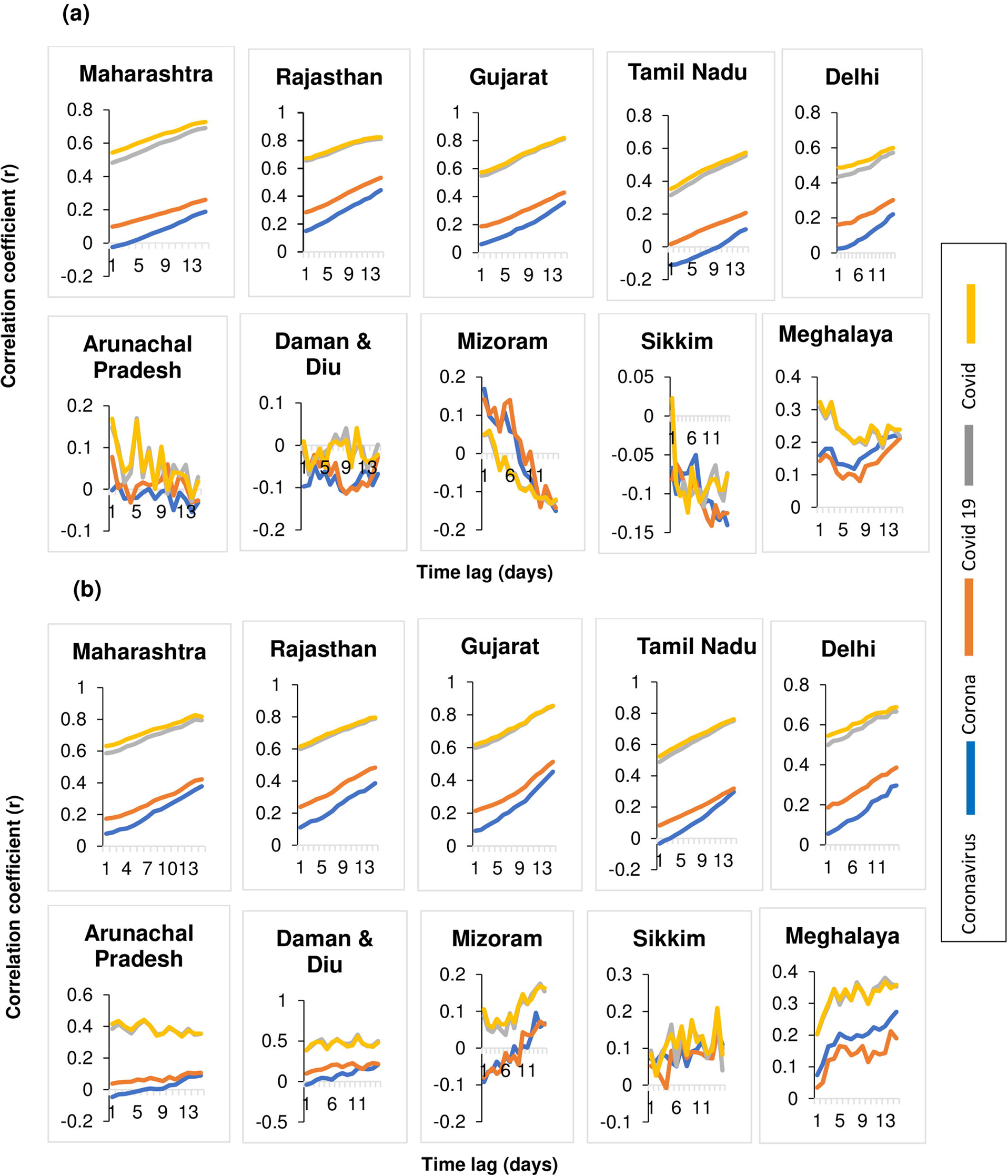
Lag correlations between Google search terms related to disease with (a) daily new laboratory-confirmed COVID-19 cases and (b) daily new tests in various states of India

**Fig 9:**
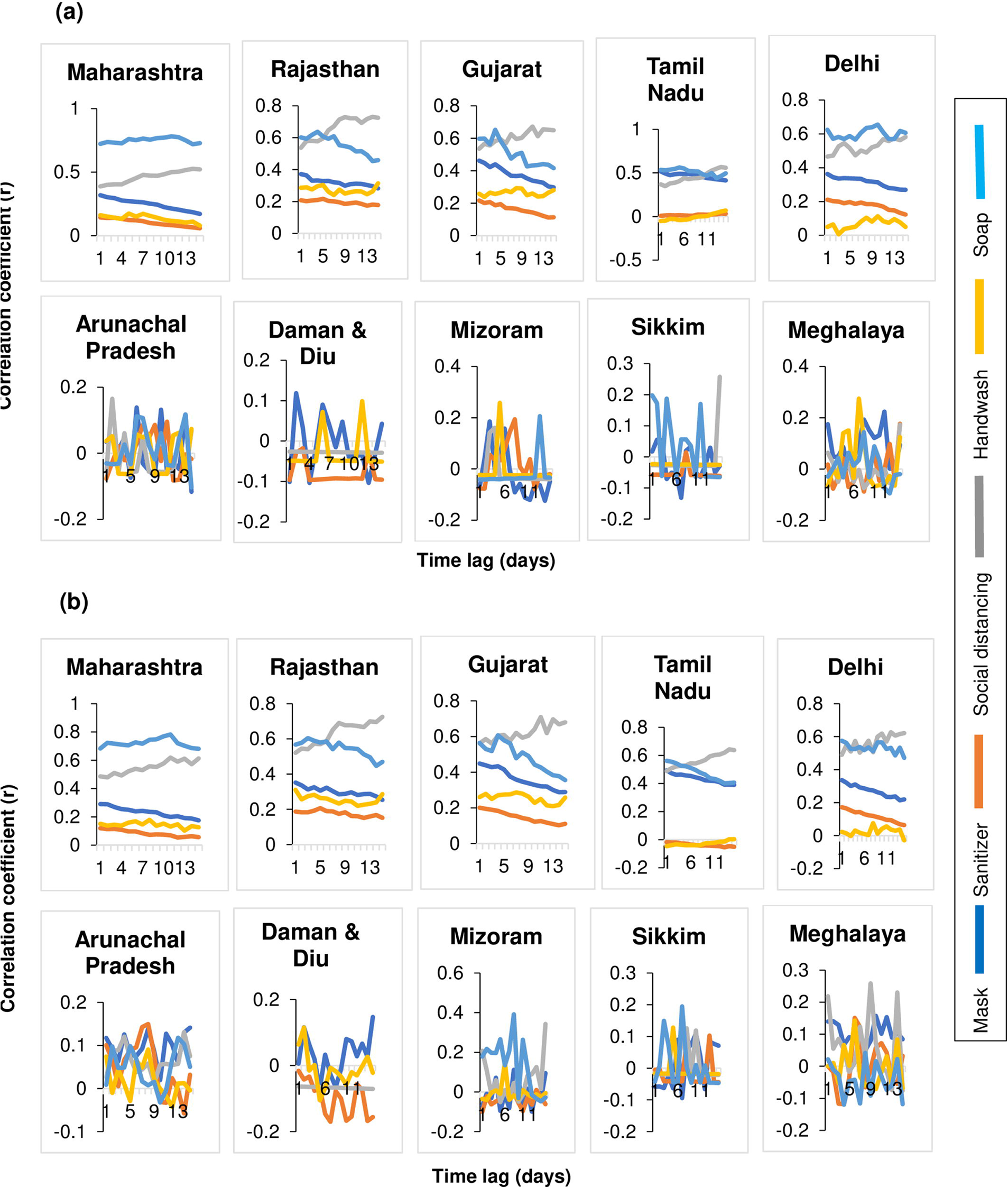
Lag correlations between Google search terms related to NPI with (a) daily new laboratory-confirmed COVID-19 cases and (b) daily new tests in various states of India

**Fig 10:**
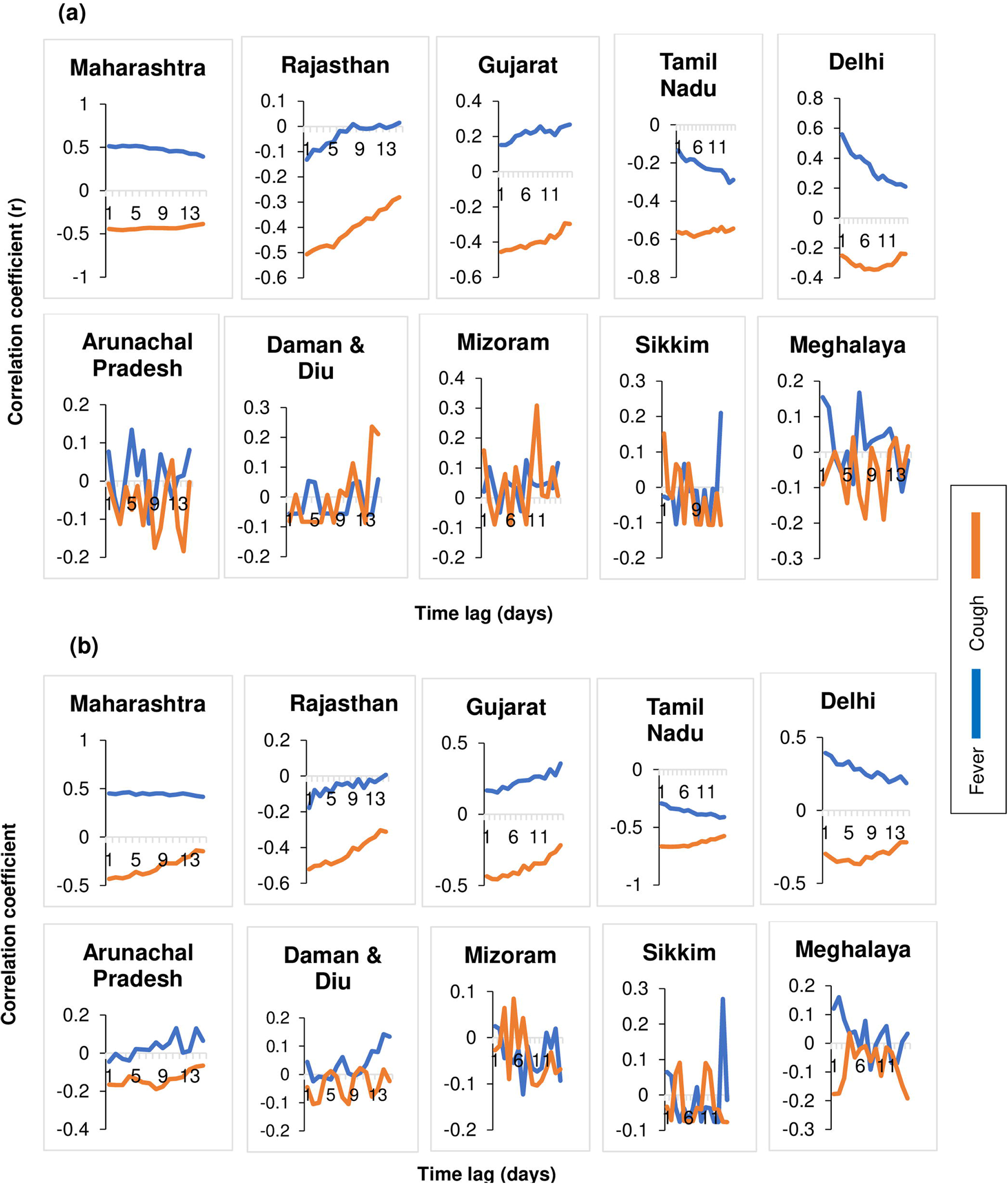
Lag correlations between Google search terms related to symptoms with (a) daily new laboratory-confirmed COVID-19 cases and (b) daily new tests in various states of India

**Fig 11:**
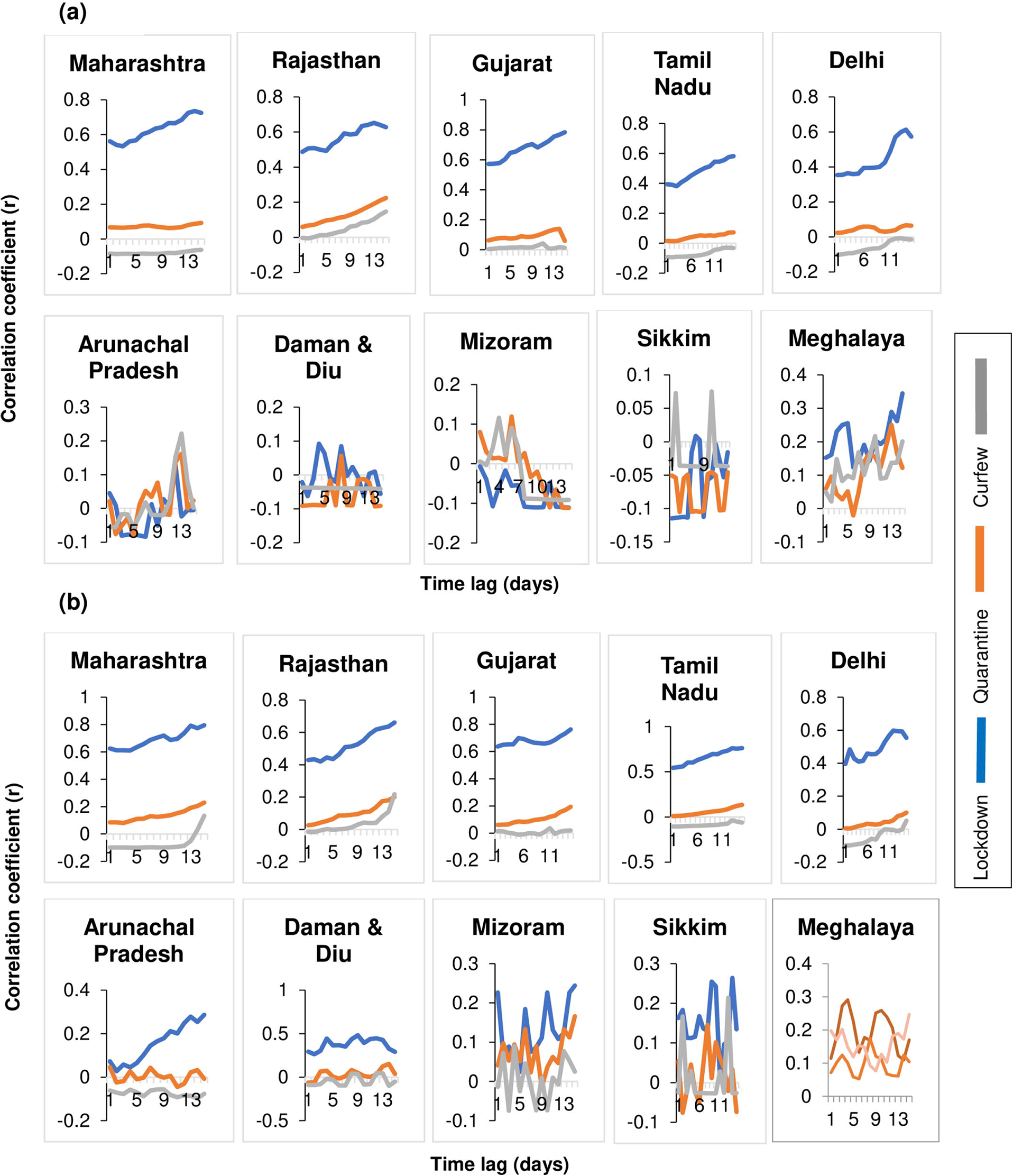
Lag correlations between Google search terms related to Government instructions with (a) daily new laboratory-confirmed COVID-19 cases and (b) daily new tests in various states of India

**Fig 12:**
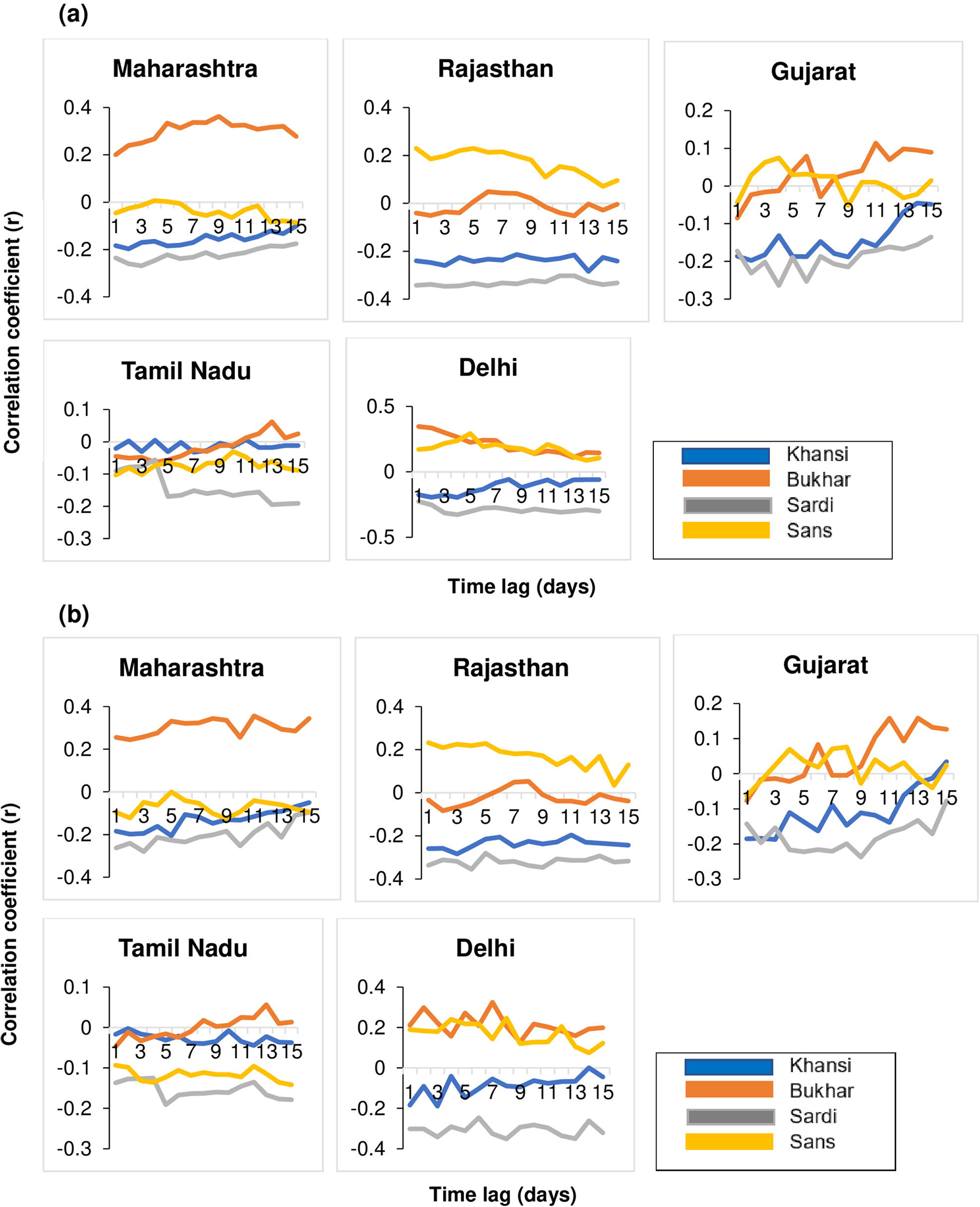
Lag correlations between Google search terms related to symptoms in Hindi (Roman script) with (a) daily new laboratory-confirmed COVID-19 cases and (b) daily new tests in various states of India

**Table 2:** Lag correlations between Google search terms related to disease and NPI and daily new laboratory-confirmed COVID-19 cases.

|  | Lag<br>(days) | Google search terms |  |  |  |  |  |  |  |  |
| --- | --- | --- | --- | --- | --- | --- | --- | --- | --- | --- |
|  |  | Coronavirus | Corona | Covid 19 | Covid | Mask | Sanitizer | Social distancing | Handwash | Soap |
| Daily new laboratory confirmed COVID-19 cases in India | 0 day | 0.017422 | 0.143727 | 0.446528 | 0.486887 | 0.348846 | 0.089318 | 0.4633163 | 0.033603 | 0.7750 |
|  | 1 day | 0.028291 | 0.154083 | 0.460124 | 0.498769 | 0.336496 | 0.088592 | 0.4656184 | 0.038185 | 0.7748 |
|  | 2 days | 0.039662 | 0.165681 | 0.471269 | 0.511705 | 0.326321 | 0.088337 | 0.4977802 | 0.040809 | 0.7749 |
|  | 3 days | 0.052734 | 0.178169 | 0.486865 | 0.525356 | 0.320705 | 0.087818 | 0.498752 | 0.041473 | 0.7768 |
|  | 4 days | 0.067589 | 0.190966 | 0.502789 | 0.542464 | 0.319026 | 0.085915 | 0.5021668 | 0.042121 | 0.7805 |
|  | 5 days | 0.085238 | 0.20719 | 0.519379 | 0.557954 | 0.312424 | 0.082179 | 0.5008109 | 0.041832 | 0.7747 |
|  | 6 days | 0.102877 | 0.220935 | 0.535928 | 0.572688 | 0.306379 | 0.079984 | 0.5050224 | 0.044593 | 0.7936 |
|  | 7 days | 0.118177 | 0.234221 | 0.550835 | 0.58665 | 0.300403 | 0.074969 | 0.5029278 | 0.047689 | 0.7919 |
|  | 8 days | 0.136251 | 0.247208 | 0.564525 | 0.600389 | 0.292124 | 0.075869 | 0.5292534 | 0.052701 | 0.8053 |
|  | 9 days | 0.153177 | 0.260884 | 0.580135 | 0.61459 | 0.28567 | 0.073306 | 0.5558849 | 0.053678 | 0.8011 |
|  | 10 days | 0.174549 | 0.275867 | 0.596239 | 0.629105 | 0.28131 | 0.072059 | 0.5869431 | 0.061314 | 0.8053 |
|  | 11 days | 0.191819 | 0.291426 | 0.613094 | 0.645633 | 0.27709 | 0.068316 | 0.5891402 | 0.049227 | 0.7951 |
|  | 12 days | 0.214608 | 0.30758 | 0.631227 | 0.66148 | 0.274595 | 0.068698 | 0.5945309 | 0.053987 | 0.7738 |
|  | 13 days | 0.234417 | 0.320818 | 0.64631 | 0.673844 | 0.266228 | 0.065646 | 0.5984586 | 0.059191 | 0.7792 |
|  | 14 days | 0.250466 | 0.33315 | 0.660226 | 0.684317 | 0.261078 | 0.064154 | 0.599263 | 0.065501 | 0.7824 |

**Table 3:** Lag correlations between Google search terms related to symptoms and government instructions with daily new laboratory-confirmed COVID-19 cases.

|  | Lag<br>(days) | Google search terms |  |  |  |  |  |  |  |  |
| --- | --- | --- | --- | --- | --- | --- | --- | --- | --- | --- |
|  |  | Fever | Cough | Lockdown | Quarantine | Curfew | Khansi | Bukhar | Sardi | Sans |
| Daily new laboratory confirmed COVID-19 cases in India | 0 day | 0.000827 | -0.57663 | 0.429016 | 0.046683 | -0.06811 | -0.51209 | -0.04085 | -0.51731 | 0.30429 |
|  | 1 day | -0.00953 | -0.57456 | 0.428641 | 0.045648 | -0.06646 | -0.50995 | -0.00744 | -0.51507 | 0.31767 |
|  | 2 days | -0.01061 | -0.57019 | 0.428088 | 0.049497 | -0.06878 | -0.49625 | -0.00601 | -0.51963 | 0.36053 |
|  | 3 days | -0.02008 | -0.56577 | 0.44224 | 0.053361 | -0.06639 | -0.49544 | 0.01564 | -0.52689 | 0.38376 |
|  | 4 days | -0.03242 | -0.55997 | 0.454573 | 0.060427 | -0.06198 | -0.49019 | 0.04372 | -0.53389 | 0.41994 |
|  | 5 days | -0.02845 | -0.55274 | 0.482757 | 0.065704 | -0.05747 | -0.47513 | 0.034626 | -0.52314 | 0.41724 |
|  | 6 days | -0.03105 | -0.54244 | 0.497414 | 0.069046 | -0.05276 | -0.46334 | 0.047201 | -0.52994 | 0.42716 |
|  | 7 days | -0.03621 | -0.53727 | 0.520351 | 0.068319 | -0.04791 | -0.44187 | 0.07466 | -0.52639 | 0.41611 |
|  | 8 days | -0.02079 | -0.52517 | 0.53134 | 0.0654 | -0.043 | -0.43511 | 0.085847 | -0.5381 | 0.46486 |
|  | 9 days | -0.02315 | -0.51392 | 0.550028 | 0.062038 | -0.04081 | -0.41341 | 0.102616 | -0.53489 | 0.48329 |
|  | 10 days | -0.0108 | -0.50685 | 0.578939 | 0.062501 | -0.03235 | -0.41232 | 0.127611 | -0.54671 | 0.48282 |
|  | 11 days | -0.00561 | -0.49144 | 0.59938 | 0.068901 | -0.02426 | -0.39921 | 0.120233 | -0.53952 | 0.48573 |
|  | 12 days | -0.00313 | -0.47494 | 0.631016 | 0.077273 | -0.01585 | -0.38184 | 0.134735 | -0.53801 | 0.44882 |
|  | 13 days | -0.00413 | -0.46206 | 0.644628 | 0.088453 | -0.0077 | -0.34684 | 0.137824 | -0.53845 | 0.47364 |
|  | 14 days | -0.00183 | -0.44549 | 0.632202 | 0.094901 | -0.00584 | -0.32527 | 0.139286 | -0.52648 | 0.44848 |

**Table 4:** Time-Lag correlations between Google search terms related to the disease and NPIs with daily new COVID-19 tests in India.

| Daily new COVID-19 tests in India | Lag (days) | Google search terms |  |  |  |  |  |  |  |  |
| --- | --- | --- | --- | --- | --- | --- | --- | --- | --- | --- |
|  |  | Coronavirus | Corona | Covid 19 | Covid | Mask | Sanitizer | Social distancing | Handwash | Soa |
|  | 0 day | 0.039334 | 0.168504 | 0.503602 | 0.543599 | 0.342228 | 0.079308 | 0.50389 | 0.033392 | 0.8111 |
|  | 1 day | 0.05123 | 0.177697 | 0.516711 | 0.556001 | 0.328896 | 0.078995 | 0.5160628 | 0.036621 | 0.8164 |
|  | 2 days | 0.061533 | 0.190811 | 0.532205 | 0.571225 | 0.31681 | 0.076506 | 0.5367087 | 0.036733 | 0.8041 |
|  | 3 days | 0.072222 | 0.204897 | 0.549907 | 0.587244 | 0.311194 | 0.073091 | 0.5312893 | 0.035037 | 0.7894 |
|  | 4 days | 0.085036 | 0.219991 | 0.566718 | 0.605578 | 0.306444 | 0.068943 | 0.5288011 | 0.033528 | 0.7741 |
|  | 5 days | 0.106059 | 0.237809 | 0.583908 | 0.620499 | 0.29748 | 0.06402 | 0.5334919 | 0.032635 | 0.7591 |
|  | 6 days | 0.129094 | 0.252702 | 0.60032 | 0.633774 | 0.287235 | 0.059209 | 0.5326655 | 0.037384 | 0.7751 |
|  | 7 days | 0.147056 | 0.266521 | 0.614188 | 0.645985 | 0.28076 | 0.053767 | 0.5443112 | 0.04246 | 0.7861 |
|  | 8 days | 0.169648 | 0.280276 | 0.628558 | 0.659706 | 0.271947 | 0.053742 | 0.5676364 | 0.047088 | 0.7931 |
|  | 9 days | 0.194573 | 0.296111 | 0.644602 | 0.675046 | 0.261912 | 0.049057 | 0.5937684 | 0.048373 | 0.7871 |
|  | 10 days | 0.215584 | 0.313861 | 0.661188 | 0.690449 | 0.258895 | 0.047572 | 0.6059841 | 0.058343 | 0.7781 |
|  | 11 days | 0.247946 | 0.331977 | 0.678266 | 0.707083 | 0.253798 | 0.04465 | 0.6162161 | 0.046952 | 0.7511 |
|  | 12 days | 0.27726 | 0.350129 | 0.695565 | 0.721583 | 0.246467 | 0.04428 | 0.6274677 | 0.053823 | 0.7331 |
|  | 13 days | 0.301974 | 0.365565 | 0.710935 | 0.733022 | 0.240909 | 0.044684 | 0.6359052 | 0.061304 | 0.7371 |
|  | 14 days | 0.31952 | 0.37896 | 0.723976 | 0.742616 | 0.238721 | 0.047759 | 0.6381407 | 0.066629 | 0.7471 |

**Table 5:** Time-Lag correlations between Google search terms related to symptoms and government instructions with daily new COVID-19 tests in India.

|  | Lag (days) | Google search terms |  |  |  |  |  |  |  |  |
| --- | --- | --- | --- | --- | --- | --- | --- | --- | --- | --- |
|  |  | Fever | Cough | Lockdown | Quarantine | Curfew | Khansi | Bukhar | Sardi | Sans |
| Daily new COVID-19 tests in India | 0 day | -0.02018 | -0.60023 | 0.490504 | 0.036242 | -0.06777 | -0.53218 | 0.000207 | -0.54266 | 0.3417 |
|  | 1 day | -0.03164 | -0.59749 | 0.487827 | 0.035859 | -0.06471 | -0.537 | 0.028383 | -0.54572 | 0.3632 |
|  | 2 days | -0.0293 | -0.59243 | 0.494749 | 0.038719 | -0.06514 | -0.51487 | 0.013564 | -0.54703 | 0.3898 |
|  | 3 days | -0.03245 | -0.5851 | 0.51395 | 0.043127 | -0.06253 | -0.50587 | 0.041926 | -0.55832 | 0.4016 |
|  | 4 days | -0.04135 | -0.57602 | 0.537027 | 0.047437 | -0.06213 | -0.49261 | 0.072069 | -0.55559 | 0.4024 |
|  | 5 days | -0.04124 | -0.56758 | 0.546561 | 0.052797 | -0.06165 | -0.47528 | 0.07654 | -0.54655 | 0.4203 |
|  | 6 days | -0.04561 | -0.55588 | 0.553328 | 0.055574 | -0.05986 | -0.45797 | 0.083631 | -0.54654 | 0.4134 |
|  | 7 days | -0.04995 | -0.54638 | 0.554689 | 0.056829 | -0.05766 | -0.43549 | 0.104467 | -0.54673 | 0.4111 |
|  | 8 days | -0.03974 | -0.53251 | 0.553329 | 0.058986 | -0.05276 | -0.42505 | 0.099752 | -0.55799 | 0.4473 |
|  | 9 days | -0.04118 | -0.51872 | 0.563061 | 0.060109 | -0.04474 | -0.39877 | 0.098395 | -0.55965 | 0.4695 |
|  | 10 days | -0.02602 | -0.50312 | 0.588455 | 0.067041 | -0.03354 | -0.3845 | 0.132634 | -0.56753 | 0.4802 |
|  | 11 days | -0.01997 | -0.48792 | 0.622881 | 0.076832 | -0.02304 | -0.36682 | 0.147165 | -0.55678 | 0.4726 |
|  | 12 days | -0.01487 | -0.46976 | 0.647796 | 0.090374 | -0.00937 | -0.34334 | 0.182885 | -0.5491 | 0.4656 |
|  | 13 days | -0.01098 | -0.45043 | 0.666029 | 0.104912 | -0.00271 | -0.30338 | 0.194457 | -0.53881 | 0.4828 |
|  | 14 days | -0.00183 | -0.42768 | 0.645629 | 0.116862 | 0.001907 | -0.27368 | 0.185472 | -0.53309 | 0.4610 |

## Trend of Disease-related terms (“coronavirus”, “corona”, “COVID 19” and “COVID”)

The first search on Google for disease related terms for both “coronavirus” and “corona” was found on 30^th^ January 2020 for India. Searches for the term “coronavirus” and “corona” began on 31^st^ January, while the first spikes happened on 4^th^ March. On this date, first search for the terms “Covid 19” and “Covid” also were observed. RSVs of 100 were observed for the terms “coronavirus”, “corona”, “COVID 19” and “COVID” on 28^th^ March, 23^rd^ March, 2^nd^ April and 28^th^ March respectively for India.

Among the states with highest caseloads, first spike for “coronavirus”, “corona” search was similar to national level (Figs 3a and 3b). The RSVs of 100 for the ten selected states were observed for the four terms on nearly the same dates except for Tamil Nadu and Meghalaya. Tamil Nadu had the RSV of 100 on 19^th^ March for “coronavirus”. RSV never reached maximum value of 31 points in Meghalaya for terms “COVID 19” and “COVID” on 14^th^ April and 15^th^ April 2020 respectively.

### Trends of Non-pharmacological Intervention (NPI) terms

The NPI terms “mask”, “sanitizer”, “social distancing”, “handwash”, and “soap” showed trends in our preliminary Google Trend search between 1^st^ January 2020 and 31^st^ May 2020. Although “mask” searches in India reached a peak on March 5^th^, but the first spike was seen on 30^th^ January 2020. The first peak coincides with the first reported case in Kerala, India. The first spikes for the term “sanitizer” was found on 5^th^ March. Afterwards, both the terms “mask” and “sanitizer” had mirrored trends with higher trend for the former term. Google searches for “social distancing”, “handwash”, and “soap” was very low throughout the observed period. (Figs 1 and 2) Google Trends™ for these terms had similar results for states/ UTs with high case load (Figs 4a and 4b). The states/ UTs with lowest caseloads didn’t have any trend, but only daily/ weekly spikes and troughs often attaining RSV of zero too. RSV reached 100 for “mask” in Arunachal Pradesh on 10^th^ March, while that for “sanitizer” in Daman & Diu (22^nd^ March), Sikkim (19^th^ March) and Meghalaya (22^nd^ March). In Mizoram, RSV reached 100 for both “mask” (9^th^ March) and “sanitizer” (18^th^ March) once in between.

### Trends for English terms for disease symptoms (“fever” and “cough”)

The terms “fever” and “cough” didn’t have distinct spikes before the peak. The RSV for the term “fever” remained more than 35 and for “cough” more than 30 throughout the reference dates. The peak search for “fever” happened on 24^th^ March 2020, while for “cough” on 23^rd^ March. Similar to Google Trends™ for disease-related and NPI terms, “fever” and “cough” had similar results for states/ UTs with high case load (Figs 5a and 5b). The states/ UTs with lowest caseloads had no clear trend, but many crest-troughs with RSV often reaching zero.

#### Trends for terms for government instructions (“lockdown”, “quarantine”, “curfew”)

The first spike of search for “lockdown” happened on 22^nd^ March 2020, followed by three more spikes on 11^th^ April, 1^st^ May and 17^th^ May which coincides with announcement of lockdown by government of India. For “quarantine” and “curfew”, there were single peaks on 22^nd^ March. Search trends for states/ UTs with highest caseloads had similar trends as national level. (Figs 6a and 6b) Among states/ UTs with lowest caseloads, Meghalaya had a single peak for the term “curfew” on 1^st^ March although the ‘junta curfew’ was declared on 21^st^ March. For Mizoram two peaks were observed on 1^st^ & 5^th^ March 2020. For the term “quarantine”, Mizoram and Daman & Diu had their peaks on 3^rd^ February and 15^th^ February 2020 respectively. Sikkim had four peaks for the term “quarantine”, first being on 9^th^ February.

### Trends for Hindi terms for disease symptoms (“khansi”, “bukhar”, “sardi”, “sans”)

The terms “khansi” and “bukhar” were also never at baseline before their peaks 21^st^ March and 22^nd^ March. Search for the term “sardi” started with a peak on 1^st^ January 2020 and then tapered to remain between RSVs of 20 and 60 between 1^st^ February and 31^st^ May 2020. The term “sans” had RSV between 20 and 50 throughout the reference dates.

States/ UTs with highest caseloads had similar trends for the Hindi terms for disease symptoms, except Delhi which had multiple peaks and troughs without any trend. (Fig 7) There was not sufficient number of searches for these terms in states/ UTs with lowest caseloads to show any trends.

### Correlation between Google Trends™ data with daily tests and cases

We determined the correlation between the 18 search terms and daily tests/ cases separately, both at national and state levels. High time lag correlation was observed between both the daily number of tests as well as daily new laboratory-confirmed cases of COVID-19 with the Google search indices for the terms “COVID 19”, “COVID”, “social distancing”, “soap” and “lockdown” at national level (Table 2). The Pearson Correlation Coefficient was highest between “soap” and Covid-19 cases reported with a time lag of 14 days (r=0.78, p<0.00001). There were some differences in the trends of the results of the time lag correlation of behavioral intervention related search terms and lockdown with daily new cases at national level. While “social distancing” and “soap” search term showed high time lag correlation with number of daily tests as well cases for the entire time lag of 14 days, the other search terms such as “hand wash”, “mask” and “lockdown” showed comparatively low time lag correlation during the period. (Table 2)

High time-lag correlation was observed between both the daily number of tests as well as daily new laboratory-confirmed cases of COVID-19 with the Google search indices for the terms “COVID 19”, “COVID”, “Corona”, “social distancing”, “soap”, “lockdown” at the level of five states with highest cases reported during the study period (Figs 8-12). In the states having lowest recorded COVID-19 cases, there was no time-lag correlation between any of the search terms and daily number of tests conducted or daily reported cases (Figs 8-12). Highest correlation of the term “soap” was seen with tests and cases in states/UTs of Maharashtra, Rajasthan, Gujarat, Tamil Nadu and Delhi. The time lag correlation between the term “soap” and number of daily tests was zero days for both Tamil Nadu and Delhi. High time-lag correlation was observed between daily new laboratory-confirmed cases of COVID-19 with the Google search indices for the term “Corona” in the state of Rajasthan at 14 days (r= 0.53, p<0.00001) while for the same term high time lag correlation with daily tests was observed at 14 days (r= 0.51, p<0.00001) in the state of Gujarat. High time-lag correlation was observed between daily new laboratory-confirmed cases of COVID-19 with the Google search indices for the term “Fever” in the states of Maharashtra and Delhi at two days (r= 0.518, p<0.00001) and zero days (r= 0.56, p<0.00001) respectively.

## Discussion

We have conducted analysis of search behaviour for Coronavirus related information through Google Trends tool for India as well as 10 states of the country with highest and lowest caseloads. We could not identify many similar studies published using similar comprehensive search strategy or including GT data from states/ UTs of India.(40) We used a comprehensive search strategy unlike many similar reported studies of Google Trends from other countries where only a few terms were used for such analysis.(25–28,30–33,40–45) Few other studies reported using additional terms through combination of literature review, clinical experience, google searches and news to compile a list of potential search terms for Google Trends analysis and used a plethora of such terms.(29,46) These studies didn’t try to categorize search terms in separate groups unlike our study. We have categorized the search terms to hypothesize the link between search trends and infodemiology outcomes for public health. Such categorization may also help in interpretation of the results in terms of search behaviour reflecting interest or apprehension and prediction of the epidemic behaviour. We argue that Google Trends analysis may be used to corroborate the interest and apprehension with disease outbreaks reported by other means of mass media (newspaper, television) and people would search internet to obtain more information for health related problems deemed having serious consequences. These search behavior may also be used to augment disease surveillance activities by providing prediction of outbreak earlier than traditional field based methods. Google Trends had shown such earlier predictive utility for H1N1 epidemic, dengue fever, Chikungunya, Malaria, etc.(13,15,47–52) As already stated India has huge number of internet users and is ranked second next only to china. (ref) Thus this use of online search behaviour is applicable for such a large userbase in India.

The duration of search for our analysis was 1st January 2020 to 31^st^ May 2020 unlike most of the other studies reporting Google Trend analysis for Covid 19 infodemiology. We chose to start our search from 1^st^ January, since the first report of novel coronavirus Pneumonia was reported from China on 31^st^ December 2019. But the first spike of search for disease-related terms happened nearly a month later. We chose 31^st^ May as the end-line of our analysis to coincide with the notification of end of lockdown in India.

The test positivity rate has remained lower in India as compared to many other countries. This might be due to lesser force of infection in India or more lenient testing guidelines. Still, there has been consistent increase in total number of cases as well as death due to COVID-19 since the detection of first case in India. But the Coronavirus disease terms related searches didn’t continue to grow in parallel with the rising number of cases. The first search is seen for the terms “coronavirus” and “corona” on 31^st^ January 2020, which reflects the media reports of first case reported on 30^th^ January. The earliest spikes on 4^th^ March related to the disease-related terms (“coronavirus”, “corona”, “COVID 19” and “COVID”) likely coincides with a cluster of cases reported among Italian tourists from Rajasthan which were reported by a section of media (print and electronic both).(53) The peak coincides with declaration of ‘junta (citizen) curfew’ and lockdown by Prime Minister of India in a televised national address on 19^th^ March and 24^th^ March 2020 respectively and associated coverage in the next one week by various sections of media.(54,55) Depending on viewership on popular television channels and newspapers, the peaks for these terms are likely explained. Similarly, the trends observed for the 10 states except Meghalaya for these four terms reflect media coverage. In the state of Meghalaya, the first case was reported on 13^th^ April, leading to the highest RSV of 31 on 14^th^ April. The heightened interest observed for the terms “coronavirus”, “corona”, “COVID 19” and “COVID” is more likely ingrained in apprehension with this unprecedented national-wide lockdown. The maximum searches related to “Covid-19” was seen in five cities of India including Mira Bhayandar (100 RSV points), Thane (87 points), Navi Mumbai (82 points), Mumbai (81 points) and Gurugram (79 points). This is likely the reflection of being places with highest caseloads and population densities. (Ref) Some other studies have also concluded that peaks in many other countries seen in Google Trends on COVID-19 disease related terms reflected media coverage than epidemic trends.(56) For example, in United States of America (USA) having highest burden of COVID-19, the peak interest was reported on 9^th^ and 12^th^ March which coincides with issuance of interim guidelines and declaration of Novel Coronavirus pandemic by World Health Organization (WHO).(24) This is in contrast to our study, where public interest was not at all present on these dates, more likely due to non-reporting of these events by mainstream media of India. For proponents of finding correlation with international events, the declaration of pandemic by Director General of WHO which happened 2 weeks prior to peak searches in India, would be chance correlation only. COVID-19-related searches in India slowly began to decline after the last week of March 2020 most likely due to flooding of information from various print, electronic and social media leading to information fatigue and disinterest. Therefore, Google Trends™ data should be used by government agencies to provide appropriate and timely risk communication to the population.(33)

We used “mask”, “sanitizer”, “social distancing”, “handwash”, and “soap” to identify search behaviour for NPI terms. Not many studies have reported this analysis in ongoing COVID-19 epidemic.(27,31,33,45) The first spike seen on 30^th^ January is likely related to the first case reported from Kerala, India. This might reflect the small group of informed people who may already be knowing that masks are one of the NPI methods of preventing respiratory illnesses like COVID-19. “Mask” related searches in India reached two peaks on March 4^th^ and 22^nd^, much earlier than the peak observed in Coronavirus related search terms. The peak on 4^th^ March likely coincides with the report of first cluster of COVID-19 cases among Italian tourists which also elicited spikes in search for “sanitizer”, “coronavirus” and “corona”. Thus, the increased search for “mask” reflects interest in protective behaviour. To verify this, we also did a search for the terms “coronavirus test” and “corona treatment”. Both these terms had their first peaks on 4^th^ March, but pushed the RSV for “mask” to nearly baseline. In an early phase of an unknown emerging disease like COVID-19, people may want to know more about methods of diagnosis and treatment rather than prevention. We hypothesize that a fraction of people who searched for test and treatment would have searched for preventive aspects like “mask”. The second peak for “mask” and “sanitizer” coincides with observation of citizen curfew on 22^nd^ March. We assume that a large population of India was confined to home on this date scouring for news as reflected by spike in television news viewership. Discussion about preventive methods might have pushed people to search for “mask” and sanitizer”, most likely for purchase. However, this search behaviour again reflects interest generated from media coverage rather than seeking health information after suffering from symptoms of respiratory illnesses. This is in contrast with similar searches in internet-savvy countries during influenza epidemics.(13) Surprisingly searches for three NPI terms “social distancing”, “handwash”, and “soap” didn’t have robust trends. These NPIs are cheaper and more likely to be correctly followed as compared to wearing masks and using sanitizers. The profile of people using internet might be quite different from the class of people who use soap as a method for prevention. The similarity in trends for NPI terms in the high caseload states/ UTs of India might be explained by their contribution to overall RSV for the country. In states with low caseloads, users were more interested in searching about sanitizer than masks in Sikkim, Meghalaya and Daman & Diu. The interest in these terms shown on 9^th^ March in Mizoram coincides with similar interest reported in USA following WHO’s interim guidelines.(24) We hypothesize local media coverage for this trend. The lesser interest in search for the symptoms (specially “fever” and “cough”) both in English and Hindi languages for COVID-19 in surprising for India. Hindi is spoken by highest number of Indians and English too is very commonly used. The few articles reporting Google Trends ™ analysis for COVID-19 didn’t use these terms.(40) Globally too, we couldn’t come across many published study which used “fever” to analyze Google Trends ™ results during ongoing COVID-19 pandemic.(29) The study by Higgins et al used a comprehensive search strategy using literature review, clinical experience, google searches and news resources. Google Trends™ analysis using the term “fever” has previously been reported for other diseases such as dengue fever and Influenza.(15,57,58) Coexistence of Influenza and COVID-19 or Dengue Fever and COVID-19 cannot be ruled out only by search behaviour using this term during winter and rainy seasons respectively. For the term “cough” too, we could not identify any published study from India using Google Trends™ analysis for surveillance. Few studies have used “cough” for Google Trends™ analysis in diseases like Sinusitis and COVID-19.(22,29,56) The peak trends for “fever” and “cough” in India coincides with the declaration of lockdown and with peak search trends for coronavirus-disease related terms and NPI terms. Similar peak trends in high burden states reflects the curiosity following declaration of lockdown. The waxing and waning of search trends for the terms “fever” and “cough” observed in low burden states/ UTs of India most likely reflects media coverage induced episodic curiosity.

Regarding trends for terms related to government instructions (“lockdown”, “quarantine” and “curfew”), peaks for “lockdown” both at national level and for the high burden states coincides with each of the five phases of lockdown from 25^th^ March to 31^st^ May 2020 enforced by government of India. The single peak for “quarantine” and “curfew” is due to the curiosity generated with observation of “junta (citizen) curfew” on 22^nd^ March. The highest trend for the term “curfew” seen on 1^st^ March in Meghalaya and Mizoram were due to local law and order issues.

We could not find any study reported from either India or other countries using Hindi terms related to COVID-19 for Google Trends™ analysis. We believe our study is the pioneer in this regard. Nearly 43.6% % of Indian population uses Hindi as their first language.(59) The term “sardi” which is the Hindi translation for both “common cold” and “winter” had its peak on 1^st^ January which coincides more aptly with the cold conditions of winter in India. This trend reflects media coverage for the extremely cold conditions in many parts of the country. The term “jukam” which more aptly is used for “common cold” in India has no trend individually, so we dropped it out of our further analysis. The terms “khansi” and bukhar” had their peaks on 21^st^ and 22^nd^ March coinciding with the observation of Junta curfew. The peak is likely explained by the curiosity emanating from media coverage of disease symptoms of COVID-19 by Hindi news channels of India throughout the day. Even in states/ UTs with highest burden of COVID-19, the trend and peak mirrored that of national level results except Delhi, more likely explained by media coverage induced interest. We are not able to explain the waxing and waning interest seen in Delhi, which is in high burden area. We hypothesize that use of these terms for Google Trends™ analysis will not help in prediction of outbreak.

This study showed high time lag correlations between the results obtained by searching for Coronavirus related keywords using Google Trends™ and the number of new daily confirmed COVID-19 cases as well as tests. Shin et al in their study on Middle East Respiratory Syndrome (MERS) using Google and twitter keywords have showed similar results with daily cases and quarantined cases.(60) These high correlations were observed maximum at 10-14 days before the confirmation of daily COVID 19 cases as well as tests. We have just explained that most of the peak for the 18 terms used for Google Trends™ analysis are explained by curiosity emanating from media coverage of the COVID-19 pandemic and events like declaration of lockdown in phases or reporting of first/ significant number of cases. Still, the positive correlation observed between RSV and numbers of daily tests or cases should be considered as supportive for its use in digital surveillance for monitoring the outbreak of COVID-19 in India. Various other studies have reported high time lag correlation between disease related search terms and the increase in cases during an outbreak.(15,25,26,28,29,33,34,60)

High time lag correlation was observed between both the daily number of tests as well as daily new laboratory-confirmed cases of COVID-19 with the Google search indices for the terms “COVID 19”, “COVID”, “social distancing”, “soap” and “lockdown” at national level. This shows increased interest among the general population not only towards COVID disease related search terms such as “COVID 19” and “COVID” but also towards their own safety as observed by the high time lag correlation throughout the period of 14 days for the terms “social distancing” and “soap”.

A high positive correlation between the number of infected cases and the Google trend values for the term “COVID 19” has been reported for India as part of analysis for eight major countries.(34) Similarly, high positive time lag correlation between the number of infected cases and the Google trend values was seen for the term “COVID” in USA.(46) While ‘Coronavirus’ term had showed high time lag correlation in several studies with cases, our study did not have such correlation. This might be due to the fact that these studies were done between January and March 2020 when ‘COVID 19’ term was not used widely.(25,28,33) Increased apprehension of the population whether the “lockdown” will end or whether it will be extended can be observed by the high time lag correlation observed with daily cases at 14 days (r = 0.63, p<0.00001) and with the daily tests at 13 days (r = 0.66, p<0.00001) at the national level in India.

“Soap” was the term with the highest Pearson correlation coefficient at a time lag of ten days (r = 0.805, p<0.00001)) when analyzed against the actual daily COVID-19 cases and a time lag of one day (r = 0.816, p<0.00001) when analyzed against the daily COVID-19 tests at the national level. This might be due to either majority of population being well informed about need and method of handwashing or not giving due importance to handwashing over use of other NPIs such as “sanitizer”. We hypothesize that media coverage of stock out of sanitizer might have prompted people to give more importance to sanitizer as compared to soap as a preventive method in COVID-19 epidemic. There were government instructions on bringing sanitizers in the list of essential commodities, which might further have deflected interest from soap to sanitizer. Moreover, government of India as well as many state government gave written instructions to shops, offices and other establishments to keep sanitizers at the entry point. Therefore, we propose that “sanitizer” may be more appropriate term for digital surveillance of COVID-19 outbreaks in India as compared to “soap” or “handwashing”. The high time lag correlation observed in the high burden states for these terms likely contributes to the observation at national level. These findings should form part of digital surveillance at state levels. It would be interesting to see whether such trend in seen in hitherto low burden states which may develop outbreaks in future. The high correlation with the term “Fever” seen in states of Maharashtra and Delhi is most likely predictive for outbreak during ongoing “COVID 19” epidemic in these states unlike at national level. This is in contrast to the results obtained by Higgins et al who have reported positive correlation between Google search for symptoms of COVID-19 and the COVID-19 cases at level of countries.(29)

These findings reflect increase in internet search activities almost ten to fourteen days before the increase in daily COVID-19 cases and deaths. These findings indicate that Google Trends™ can be used as a tool to monitor public restlessness toward COVID-19 infections in India as well as potentially define the proper timing as well as location of risk communications. The utilization of web-based data for health care research, otherwise called “infodemiology”, is a promising new domain and it can very well complement the existing data sources.(61) Providing accurate and early information during an outbreak in the form of risk communication is the need of the hour. Appropriate risk communication can help avoid “infodemics” which might induce panic and apprehension among the population and leading to inappropriate utilization of scarce health resources. As there is little knowledge associated with emergence of newer diseases such as Novel Coronavirus, this availability of prompt, accurate and early information for monitoring the infectious disease will be worth its weight to make informed decisions for controlling the disease. The internet has become an important source of heath information across the globe more so in times of outbreak of infectious diseases. Previous epidemics have supported the use of internet searches for digital outbreak surveillance, pointing towards the fact that the digital surveillance deserves greater investment by the public health agencies.(62–64) Various government agencies like MoHFW, PMO, Health Ministry, ICMR,PIB, MyGov portal etc have shared risk communication and education material on COVID-19. But the viewership of these portal is lower as compared to other private media platforms (print and electronic). Therefore, the risk communication models by the government should incorporate these privately owned media platforms to provide appropriate health information. It was observed in a study conducted to evaluate Google Trends™ to determine its reliability as a tool for epidemiology that it is definitely influenced by media coverage.(65)

Data from tools such as Google Trends™ may predict the actual disease outbreak much earlier than the routine surveillance by the government agencies because a majority of population use web based data to access health information related to outbreak of any infectious disease. The advantages of using a tool such as Google Trends™ is that data can be obtained earlier, more easily and at a much lower cost than routine surveillance techniques adopted by the governments. They should at least be used to supplement the surveillance data obtained by traditional methods. A study has compared the surveillance platform of India known as Integrated Disease Surveillance Project (IDSP) and reported strong correlation with a time lag of 2-3 weeks for Chikungunya, Dengue Fever, Malaria and Enteric Fever.(50) Similar studies should be attempted for COVID-19 data from IDSP and Google Trend™. We also recommend that longitudinal studies be conducted among users of internet who have used health-related search to find the reason(s) for such searches. The results of such studies will corroborate whether Google Trends™ can be used as a proxy for illnesses among general population. Correction factors on internet usage behaviour may be used to the illness-induced Google search for health information and supplement or complement traditional surveillance system in India. We hypothesize that lack of such confirmatory studies led to demolishing of Google Flu search network.(66)

Our study does have certain limitations. The selection of the keyword as well as the associated spelling might affect the overall results of the study. There is no globally accepted guideline for Google Trends™ analysis, though a procedure has been recommended by some authors.(61) A guideline is required from Google™ in this regard who has custody of the search data, but doesn’t share the algorithm for search.(67)

## Conclusion

This study reveals the advantages of infodemiology using Google Trends™ to monitor an emerging infectious disease like COVID-19 in India. Reliable surveillance data can be obtained earlier, more easily and at a lower cost than traditional surveillance. The results of this study show that Google Trends™ data are highly correlated with COVID-19 tests and cases in India. Hence this platform can be used to increase the preparedness and responsiveness at various regional levels, which is paramount in handling the current pandemic of COVID-19.

## Data Availability

We obtained data on cases of COVID-19 in India at country level from WHO COVID-19 dashboard.
We obtained data on numbers of tests (both country and state level) and cases (state-level) from covidindia.org website.
We obtained the google search trend data from Google Trends ™ website.

https://covid19.who.int

https://www.covid19india.org

https://trends.google.com/trends/?geo=US

## Acknowledgements

We are thankful to Alphabet Inc™ which provides access to use Google Trends™ platform. We are thankful to WHO and ECDC which provides free access to COVID-19 global database. We also thank developers of the website covid19india.org which provides dynamic tracking of COVID-19 related testing, cases and deaths statistics for India. We are also thankful to PIB which provides periodic information on various government related communication, specifically for COVID-19 related updates.

## Data Availability Statement

All relevant data are within the paper and its Supporting Information files.

## Supporting Information

**S1 File. Search strategy**

**S2 File. Dataset**

## Author Contributions

**Conceptualization: PS, SK, PP**

**Data curation: PS**

**Formal analysis: PS**

**Methodology: PS, SK, PP**

**Project administration: Not applicable**

**Resources: PS, SK, PP**

**Software: PS**

**Supervision: SK**

**Validation: PS, SK, PP**

**Visualization: PS**

**Writing – original draft: SK**

**Writing – review & editing: PS, SK, PP**

